# The Australian Genomics Mitochondrial Flagship: A National Program Delivering Mitochondrial Diagnoses

**DOI:** 10.1101/2023.11.08.23298009

**Authors:** Rocio Rius, Alison G. Compton, Naomi L. Baker, Shanti Balasubramaniam, Stephanie Best, Kaustuv Bhattacharya, Kirsten Boggs, Tiffany Boughtwood, Jeffrey Braithwaite, Drago Bratkovic, Alessandra Bray, Marie-Jo Brion, Jo Burke, Sarah Casauria, Belinda Chong, David Coman, Shannon Cowie, Mark Cowley, Michelle G. de Silva, Martin B. Delatycki, Samantha Edwards, Carolyn Ellaway, Michael C. Fahey, Keri Finlay, Janice Fletcher, Leah E. Frajman, Ann E. Frazier, Velimir Gayevskiy, Roula Ghaoui, Himanshu Goel, Ilias Goranitis, Matilda Haas, Daniella H. Hock, Denise Howting, Matilda R. Jackson, Maina P. Kava, Madonna Kemp, Sarah King-Smith, Nicole J. Lake, Phillipa J. Lamont, Joy Lee, Janet C. Long, Mandi MacShane, Evanthia O. Madelli, Ellenore M. Martin, Justine E. Marum, Tessa Mattiske, Jim McGill, Alejandro Metke, Sean Murray, Julie Panetta, Liza K. Phillips, Michael C.J. Quinn, Michael T. Ryan, Sarah Schenscher, Cas Simons, Nicholas Smith, David A. Stroud, Michel C. Tchan, Melanie Tom, Mathew Wallis, Tyson L. Ware, AnneMarie E. Welch, Christine Wools, You Wu, John Christodoulou, David R. Thorburn

**Affiliations:** Centre for Population Genomics, Murdoch Children’s Research Institute, Melbourne, VIC, Australia; Centre for Population Genomics, Garvan Institute of Medical Research, and UNSW Sydney, Sydney, NSW, Australia; The University of Melbourne, Melbourne, Australia; Murdoch Children’s Research Institute, Melbourne, Australia; Victorian Clinical Genetics Services, Melbourne, Australia; Sydney Children’s Hospitals Network, Westmead, Australia; University of Sydney, Sydney, NSW, Australia; Australian Institute of Health Innovation, Macquarie University, Sydney, Australia; Peter MacCallum Cancer Centre, Melbourne, VIC, Australia; Victorian Comprehensive Cancer Centre, Melbourne, VIC, Australia; Australian Genomics, Murdoch Children’s Research Institute, Melbourne, VIC, Australia; Women’s and Children’s Hospital, Adelaide, Australia; QIMR Berghofer Medical Research Institute, Brisbane, Australia; Tasmanian Clinical Genetics Service, Hobart, Australia; The University of Tasmania, Hobart, Australia; Queensland Children’s Hospital, Brisbane, Australia; Wesley Hospital, Brisbane, Australia; University of Queensland, Brisbane, Australia; Children’s Cancer Institute, University of New South Wales, NSW, Australia; Harry Perkins Institute of Medical Research, University of Western Australia, Perth, Australia; University of Sydney, Sydney Australia; Royal Melbourne Hospital, Melbourne, VIC, Australia; Royal Adelaide Hospital, Adelaide, Australia; Garvan Institute of Medical Research, Sydney, NSW, Australia; John Hunter Hospital, Newcastle, Australia; Bio 21 Molecular Science and Biotechnology Institute, Melbourne, Australia; Harry Perkins Institute of Medical Research, Perth, Australia; Department of Genetics and Molecular Pathology, SA Pathology, Adelaide, Australia; Perth Children’s Hospital, Perth, Australia; The Australian e-Health Research Centre, CSIRO, Australia; Yale School of Medicine, New Haven, CT, USA; Royal Perth Hospital, Perth, Australia; Royal Children’s Hospital, Melbourne, Australia; Mito Foundation, Sydney, Australia; Royal Melbourne Hospital, Melbourne, Australia; Mater Hospital, South Brisbane, Australia; Australian Genomics, Genetic Health Queensland, Brisbane, Australia; Monash University, Melbourne, Australia; Department of Neurology and Clinical Neurophysiology, Women’s and Children’s Hospital, Adelaide, South Australia, Australia; Discipline of Paediatrics, University of Adelaide, Adelaide, South Australia, Australia; Faculty of Medicine and Health, University of Sydney, Australia; Department of Genetic Medicine, Westmead Hospital, Westmead, Australia; Genetic Health Queensland, Brisbane, Australia; School of Medicine and Menzies Institute for Medical Research, University of Tasmania, Hobart, Australia; Royal Hobart Hospital, Hobart, Australia

## Abstract

**Purpose:** Families living with mitochondrial diseases (MD) often endure prolonged diagnostic journeys and invasive testing, yet many remain without a molecular diagnosis. The Australian Genomics Mitochondrial flagship, comprising clinicians, diagnostic, and research scientists, conducted a prospective national study to identify the diagnostic utility of singleton genomic sequencing using blood samples.

**Methods:** 140 children and adults living with suspected MD were recruited using modified Nijmegen criteria (MNC) and randomized to either exome + mtDNA sequencing (ES+mtDNAseq) or genome sequencing (GS).

**Results:** Diagnostic yield was 55% (n=77) with variants in nuclear (n=37) and mtDNA (n=18) MD genes, as well as phenocopy genes (n=22). A nuclear gene etiology was identified in 77% of diagnoses, irrespective of disease onset. Diagnostic rates were higher in pediatric-onset (71%) than adult-onset (31%) cases, and comparable in children with non-European (78%) versus European (67%) ancestry. For children, higher MNC scores correlated with increased diagnostic yield and fewer diagnoses in phenocopy genes. Additionally, three adult patients had a mtDNA deletion discovered in skeletal muscle that was not initially identified in blood.

**Conclusion:** Genomic sequencing from blood can simplify the diagnostic pathway for individuals living with suspected MD, especially those with childhood onset diseases and high MNC scores.

## A) INTRODUCTION

Mitochondrial diseases (MD) are a heterogeneous group of disorders caused by pathogenic variants in nearly 400 genes leading to mitochondrial dysfunction and impaired ability of cellular energy generation.^1^ The phenotypic spectrum of MD is very broad and can affect many different organs, including the brain, heart, muscles, and the nervous system. They are the most common group of inherited metabolic disorders with a prevalence of at least 1 in 5,000 live births.^2,3^ This group represents the highest mortality in the pediatric population among all inherited metabolic disorders.^4^

Identification of an underlying molecular diagnosis for patients and their families living with suspected MD is crucial for informing clinical management, gaining insight about prognosis, and allowing families to make informed reproductive decisions. These diagnoses can also facilitate further mechanistic research, which may ultimately lead to the development of novel treatments.

The diagnosis of MD has traditionally been based on a combination of clinical criteria, along with biochemical and genetic testing, which often varies depending on the clinical presentation. However, the complexity and variability of these diseases has made it challenging to accurately diagnose them due to various factors, including the fact that they can be caused by pathogenic variants in either nuclear DNA (nDNA) or mitochondrial DNA (mtDNA), the different modes of inheritance, phenotypic variability, and the levels of mtDNA heteroplasmy. Often families with MD have visited multiple specialists, been misdiagnosed, or required extensive evaluations, including biopsies to perform biochemical, histological, and enzyme evaluations.^5^

Exome (ES) and genome (GS) sequencing have emerged as powerful tools for diagnosing MD. These sequencing technologies allow for the simultaneous testing of multiple genes and have improved the diagnostic yield and the identification of novel disease genes.^6,7^

ES has gained widespread adoption due to its lower cost and ability to target nearly all coding regions and flanking intronic nucleotides. To screen for mtDNA variants, “off-target” reads from ES can be analyzed. However, this method depends on the specific exome kit employed as the entire mtDNA is often not equally captured and can be limited in its ability to detect and accurately quantify low levels of heteroplasmy, as the sequencing depth may be variable.^8,9^ Alternatively, additional targeted mtDNA sequencing (mtDNAseq) could be performed.^10,11^ Genome Sequencing (GS) can comprehensively interrogate both nDNA and mtDNA in a single test detecting variants in the coding and non-coding regions of the genome.^6^

For individuals with suspected MD, using blood samples for genomic testing (GS/ES+/- mtDNAseq) is particularly attractive as it could also potentially obviate the need for invasive testing. The diagnostic yield of GS and ES in the context of MD ranges from 31% to 70%.^12,13^ The variability in diagnostic yields may be due to differences in the stringency of inclusion criteria, previous testing, study design, and the inherent heterogeneity among patient cohorts. To better understand the clinical diagnostic utility of these technologies in individuals living with MD, Australian Genomics established the *Mitochondrial flagship*, assembling a national team of clinicians, diagnostic, and research scientists who conducted a prospectively designed study by selecting children and adults living with suspected MD using modified Nijmegen criteria (MNC) (Table S1)^6,14^ and randomized for testing through ES+mtDNAseq or GS using DNA extracted from blood as a first step.

## B) MATERIALS AND METHODS

### Study participants

Prospectively identified individuals with a “probable” (score 5–7) or “definite” (score 8–12) diagnosis of MD based on MNC were eligible for recruitment.^14^ Thirteen individuals with a score of 4 (“possible diagnosis”) and without a previous muscle biopsy were accepted because there was consensus by an expert clinical intake committee warranting investigation. The expert clinical intake committee recognized that the absence of a muscle biopsy likely limited their ability to achieve a higher score under the MNC, which allocates points for biopsy results. The committee agreed to prioritize non-invasive methods where feasible and by consensus determined that their clinical presentations suggested probable mitochondrial disease, warranting further investigation. Patients were excluded if they had a previously confirmed molecular diagnosis, previous testing through ES or GS, or an indication that there is another likely non-MD diagnosis from other investigations as determined by the intake review committee.

A total of 140 individuals were recruited between 2017-2020 from the states of New South Wales (n=29), Queensland (n=40), South Australia (n=12), Victoria (n=40), Tasmania (n=2) and Western Australia (n=17). Individuals were randomized to be studied through singleton ES+mtDNAseq or singleton GS using DNA extracted from blood as a first step. Patients were classified based on the age of onset of their symptoms. Pediatric-onset patients included those who developed symptoms before the age of 16, including 20 individuals who continue to be followed into adulthood. Adult-onset patients comprised those who developed symptoms at or after 16 years of age.

### Genetic analysis

The genetic analysis iteratively developed over the course of the study (Figure S1). Initially individuals underwent GS or ES+mtDNAseq from samples in blood as follows:

### Exome and mitochondrial DNA sequencing

ES was performed at the Victorian Clinical Genetics Services (VCGS) using the Agilent SureSelect^QXT^ CREv1 and CREv2 kit on Illumina sequencing instruments, with a targeted mean coverage of 100x and a minimum of 90% of bases sequenced to at least 15x. Data were processed using Cpipe^15^ to generate annotated variant calls within the target region (coding exons +/- 2bp), via alignment to the reference genome (GRCh37). SNV analysis in the ES cohort was performed using an in-house analysis pipeline. CNV analysis from exome sequencing data was performed in selected individuals using an internal tool CxGo^16^ when a gene of interest was identified.

mtDNAseq was performed if initial ES analysis was negative (n=59/72). For mtDNAseq, the whole mitochondrial DNA (mtDNA) of 16.5kb was amplified with a single long-range PCR, followed by Illumina Nextera^®^ XT library preparation and sequencing on a MiSeq using v2 chemistry at VCGS^10,11^ with a minimum coverage of 1000-fold. Raw sequencing data were analyzed with MiSeq Reporter (v2-5-1), which was used to align sequencing reads to the revised Cambridge Reference Sequence (rCRS) mitochondrial genome (NC_012920.1) and to generate both BAM and VCF files, as well as assay quality metrics. A custom in-house analysis pipeline was used to annotate the VCF file with variant information, which was used to perform variant filtration and prioritization. This assay is clinically validated to detect SNVs with heteroplasmy >3%. The BAM file was used to generate coverage and split read plots for detection of large (>1kb) deletions.

### Genome sequencing

TruSeq Nano libraries were prepared and loaded onto a HiSeq X Ten sequencer (Illumina; control Software v3.0.29.0) and 2 x 150 bp paired-end sequencing was performed at the Kinghorn Centre for Clinical Genomics (KCCG). Raw sequencing data were converted to FASTQ format using Illumina’s bcl2fastq converter (v2.15.0.4) and read quality was evaluated using FASTQC. Sequences were aligned to the b37d5 human reference genome using Burrows-Wheeler Aligner (BWA, v0.7.12-r1039), coordinate-sorted using Novosort (v1.03.04, Novocraft Technologies Sdn Bhd, Selangor, Malaysia), and improved using GATK (v3.4-46-gbc02625) indel realignment and base recalibration to generate BAM files. Variants were called using GATK (v3.4-46-gbc02625) HaplotypeCaller followed by joint variant calling with GenotypeGVCFs and VariantRecalibration.^17^ The resulting multi-sample VCF file was annotated using ENSEMBL’s Variant Effect Predictor (v74) and converted to an SQLite database using gemini (v0.17.2).^18^ Gemini databases were imported into Seave^19^ which was used to perform variant filtration and prioritization for initial GS analyses.

Mitochondrial SNV and indels were identified using mity^20^ optimized to identify low heteroplasmy variants (<1%), with an average coverage of 3000-fold of the mitochondrial genome. Structural variants (SV) including copy number variants (CNVs) were investigated using ClinSV.^21^

### Updated Genome and Exome analysis

Expanded analyses of the GS and ES data were performed using updated pipelines at the Centre for Population Genomics; in brief the reads were realigned to the UCSC GRCh38/hg38 reference genome using Dragmap (v1.3.0). Cohort-wide joint calling of single nucleotide variants (SNVs) and small insertion/deletion (indel) variants was performed using GATK HaplotypeCaller (v4.1.4.1) with “dragen-mode” enabled.^17^ Variants were annotated using VEP (v105) and loaded into the web-based variant filtration platform, Seqr.^22^ Sex was inferred from the genotypes using the Somalier tool.^23^

Variant filtration and prioritization were performed using gene lists from PanelApp (Australia)^24^, initially using *mitochondrial diseases* (Version 0.850) and *mendeliome* (Version 1.571) gene lists. If a diagnosis was not reached, an expanded analysis was performed using a custom *mitoexome* gene list, which includes genes related to mitochondrial function (Table S2). Variant curation was based on the American College of Medical Genetics and Genomics (ACMG) guidelines^25^, and Variants of Uncertain Significance (VUS) were further sub-classified as being of potential clinical relevance (class 3A), uncertain significance (class 3B) or with low clinical relevance (class 3C).Reanalysis was performed pragmatically and triggered by technological advancements, data realignment to GRCh38/hg38, transitions between analysis platforms (e.g., Seave to Seqr), gene list updates, and receiving new clinical information.

## C) RESULTS

One hundred and forty individuals were recruited into this study (85 pediatric and 55 with adult-onset), and the characteristics for each sequencing arm are summarized in Table 1 and Table S3. Parental self-reported ancestry was recorded by clinicians or genetic counsellors and classified according to Human Ancestry Ontology and Australian Standard Classification of Cultural and Ethnic Groups.^26^ Notably, the majority (78%) of the adult-onset group identified as European. This demographic pattern differed in the pediatric-onset group, which showed more diversity. Here, only 46% of participants had both parents self-identifying as “European” (Figure 1A). This demographic shift can be contextualized by considering Australia’s history of successive waves of migration.^27^ Additionally, consanguinity was reported in 1 family of European ancestry and 8 with non-European ancestry.

**Figure 1.**
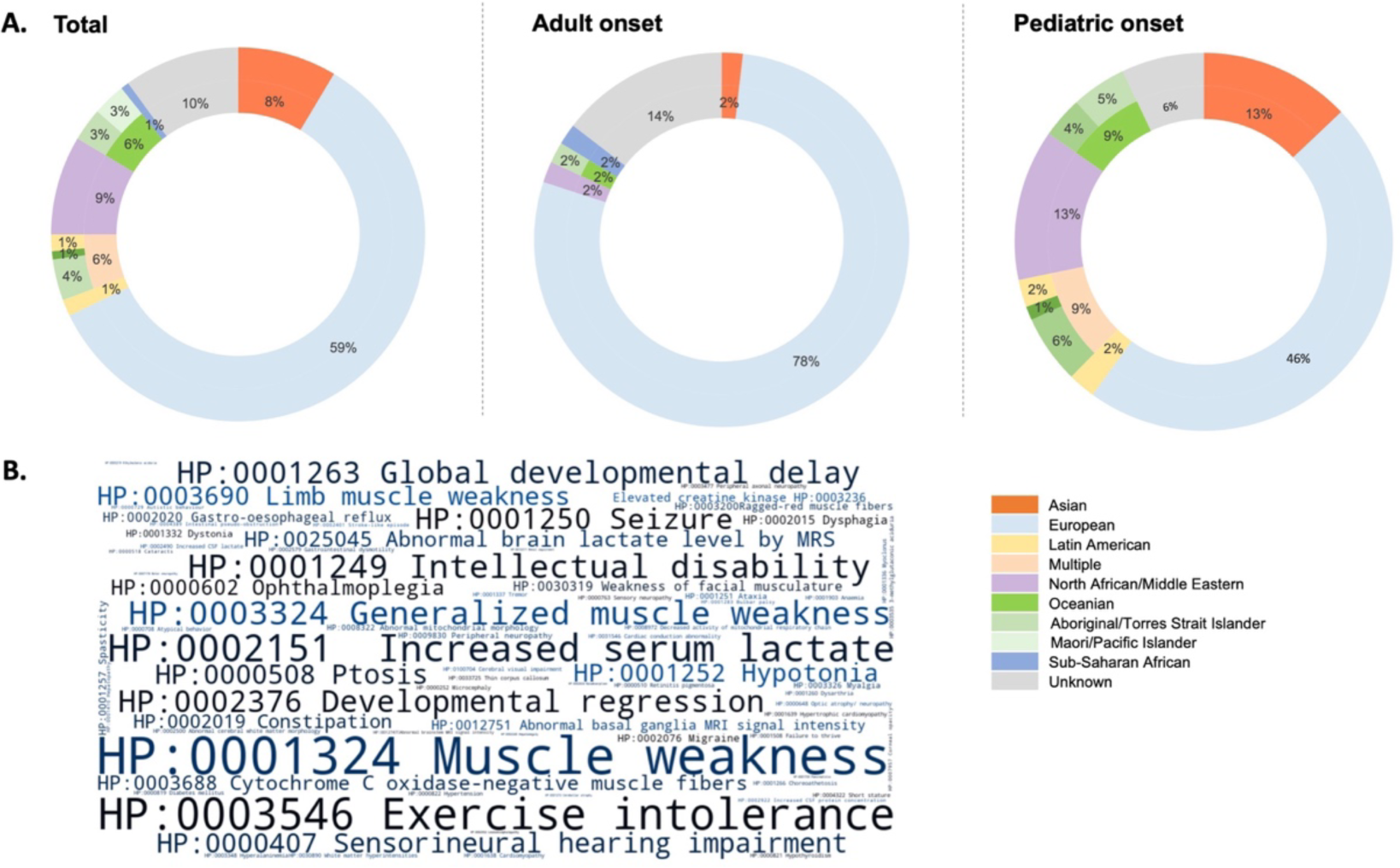
Self-reported ancestry and Human Phenotype Ontology (HPO) Term Frequencies observed in the cohort. A. Self-reported ancestry in the total cohort and by age of onset groups. B. Word Cloud of HPO terms, size, and darkness of each term within the cloud represent its prevalence within the cohort, illustrating the most common phenotypes.

**Table 1.** Mitochondrial Flagship individual characteristics.

| Characteristic | All individuals | ES +mtDNAseq | GS | ES+mtDNAseq<br>vs GS<br>p value |
| --- | --- | --- | --- | --- |
| **Sex (n, %)** |  |  |  | 0.608 |
| Male | 57 (41%) | 31 (43%) | 26 (38%) |  |
| Female | 83 (59%) | 41 (57%) | 42 (62%) |  |
| **Age of onset (n, %)** |  |  |  | 0.7315 |
| Adult | 55 (39%) | 29 (40%) | 26 (38%) |  |
| Pediatric | 85 (61%) | 43 (60%) | 42 (62%) |  |
| **Modified Nijmegen**<br>**score**<br>**(median, IQR)** | 6 (5 - 7) | 6 (5 - 7) | 6 (5 - 7) | 0.7591 |

IQR denotes interquartile range. Pediatric individuals were younger than 16 years of age at onset of clinical symptoms.

Human Phenotype Ontology (HPO) terms were extracted from the phenotypic data entries using the CSIRO Fast Healthcare Interoperability Resources terminology server^28^ and by manually inspecting intake forms and clinical summaries. In the cohort, each individual exhibited a range of 4 to 23 distinct HPO terms, resulting in a total of 121 unique entries with a combined total of 1503 occurrences across the cohort. The most frequent terms were HP:0001324 muscle weakness (n=81, 58%), HP:0003546 exercise intolerance (n=75, 54%), HP:0002151 increased serum lactate (n=75, 54%), HP:0001249 intellectual disability (n=54, 39%), HP:0001263 global developmental delay (n=47, 34%), HP:0002376 developmental regression (n=47, 34%), HP:0001250 seizure (n=43, 31%), HP:0000407 sensorineural hearing impairment (n=40, 29%), HP:0000508 ptosis (n=39, 28%) (Figure 1B).

A likely molecular diagnosis was identified in 55% of individuals in the cohort (n=77), including seven individuals with “strong candidate” diagnoses in known disease genes, one with a novel disease gene association (*UNC13A*), and one with a phenotypic expansion (*TOP3A*). Ongoing work, including functional studies, is being conducted to confirm their causality. Seventy one percent (n= 55) of the total cohort diagnoses were in known MD genes, of which 67% (n=37) were nuclear and 33% (n=18) mitochondrial genome in origin (Figure 2). For 29% (n=22) of the diagnoses, the causative genes were not known to have a mitochondrial function (i.e., a phenocopy). Most of the diagnoses were due to single nucleotide variants (SNV) (n=68); other types of variants included three duplications involving the *ATAD3* gene cluster (*ATAD3A*; HGNC:25567, *ATAD3B*; HGNC:24007, *ATAD3C*; HGNC:32151) ^29^, four single large mtDNA deletions, and intragenic deletions that were identified *in trans* with a SNV in two individuals (P3-*SERAC1,* HGNC:21061; P135 – *AARS2,* HGNC:21022).

**Figure 2.**
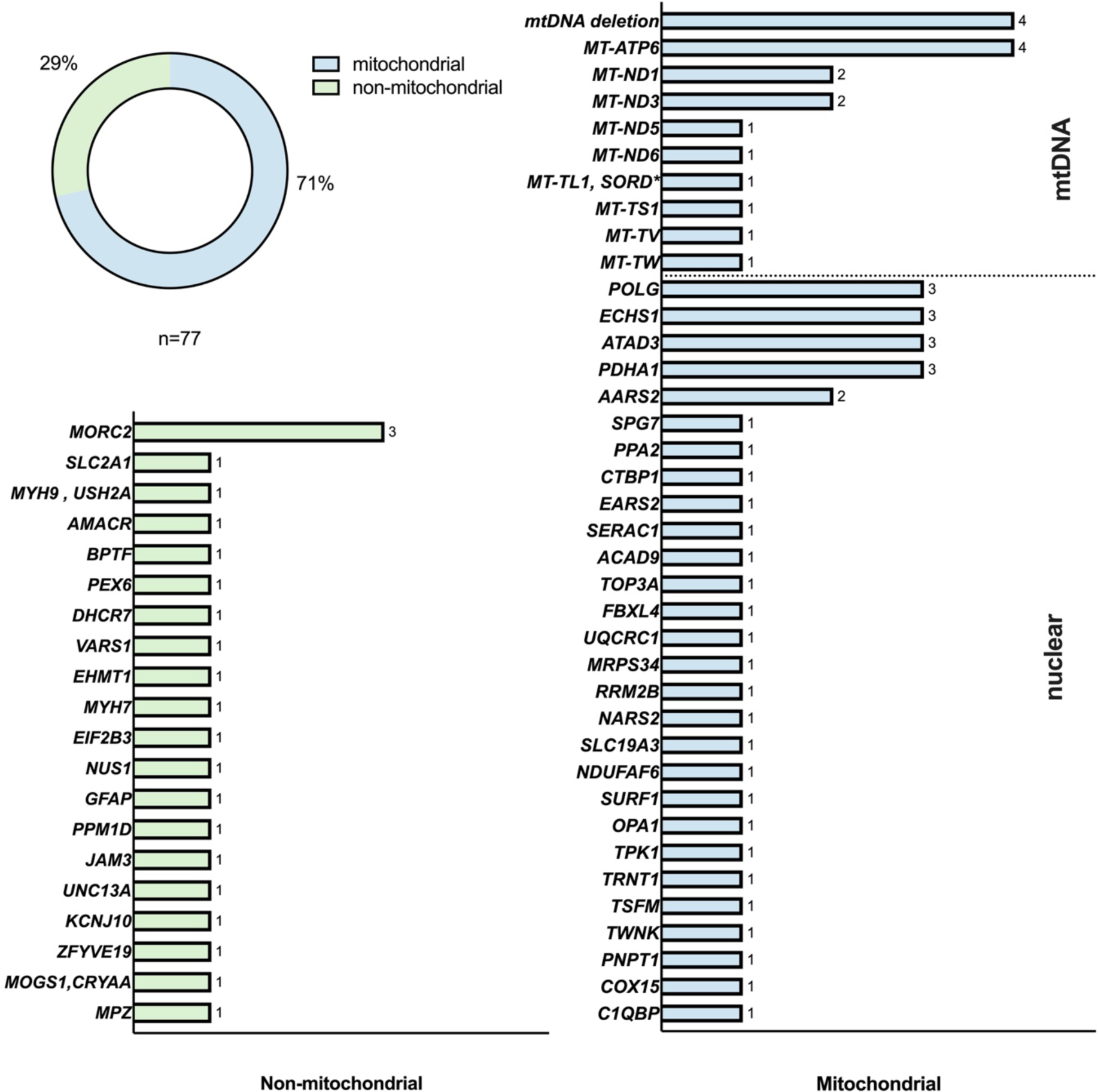
Number of individuals with a likely molecular diagnosis in mitochondrial and non-mitochondrial disease genes. \**SORD* is not a mitochondrial disease gene; however, the individual is listed in the mtDNA group as most of the phenotype is explained by the *MT-TL1* pathogenic variant.

Three individuals had a dual diagnosis, two in non-mitochondrial disease genes and one with a MD and non-mitochondrial disorder. The dual non-mitochondrial disease diagnoses were in *MOGS1* (HGNC:24862) and *CRYAA* (HGNC:2388) in individual P5^30^, and in *MYH9* (HGNC:7579) and *USH2A* (HGNC:12601) in individual P119^31^. Individual P47 had pathogenic variants in the mtDNA encoded MD gene *MT-TL1* (HGNC:7490), explaining most of his symptoms, and in *SORD* (HGNC:11184; a non-mitochondrial disease gene) contributing to some of the phenotype. One additional individual, P117, had a partial diagnosis identified in *MYH7* (HGNC:7577).

Six individuals had diagnostic SNVs identified through mtDNA sequencing in blood after initial clinical WES analysis. Three of these SNVs, located in protein-coding mtDNA genes, were detectable upon reanalysis of WES data from off-target reads (Table S4). A heteroplasmic variant (22%) in MT-ND3 was not detected, and two variants in mt-tRNA genes were missed, as exome capture kits often do not target these genes. Due to the potential for false positives from nuclear mitochondrial DNA segments (NuMTs), it is recommended to confirm mtDNA variants identified from off-target WES using an alternative method.

During the first stage of analysis, the GS data were interrogated for variants in coding regions of known disease genes (*mitochondrial disease* and *Mendeliome* gene lists). Given this focus on coding regions, it is plausible to assume that if the exome sequencing had robust coverage of these regions, most of the SNVs in the GS arm could have been identified by ES+mtDNAseq. In the expanded GS, non-coding regions of known and candidate disease genes were interrogated, which was not technically possible in the ES+mtDNAseq group.

We subsequently performed secondary GS in 14 individuals from the ES+mtDNAseq cohort where there was a high diagnostic suspicion (such as a single variant of interest in a gene associated with an autosomal recessive disease) and where DNA was available. This resulted in an additional probable diagnosis in P67 as secondary GS identified a deep intronic “second hit” NM_003365.3:c.707-186G>A (SpliceAI donor gain 0.23) in *UQCRC1* (HGNC:12585), currently undergoing functional studies.

In addition, during the expanded analysis, muscle mtDNA testing was suggested if the phenotype was compatible with a mtDNA deletion or a low heteroplasmy variant was identified in blood. Of the 12 individuals who had mtDNAseq or alternate genetic testing (such as southern blotting) in muscle after enrolling in the study, one adult was confirmed to have higher heteroplasmy levels of a pathogenic SNV in *MT-TL1* (HGNC:7490) first identified in blood (m.3243A>T 2% in blood; 69% in muscle), and a single large mtDNA deletion was identified in muscle from three individuals (P8, P56 and P124).

The diagnostic rate of individuals who started the diagnostic pathway with GS from blood was 56% (n=38). In one individual from this arm, the molecular diagnosis was identified through mtDNAseq in muscle (P56) following non-diagnostic GS (Figure S2). The diagnostic yield of individuals who started their diagnostic trajectory with ES+mtDNAseq was 54% (n=39). However, two individuals were diagnosed with a mtDNA deletion in muscle that was not initially identified in blood (P8, P124) (Figure S2), and in one individual, the presumed molecular diagnosis was achieved after secondary GS (P67). After excluding these four individuals, the diagnostic yield was 54% (n=37) for GS and 50% (n=36) for ES+mtDNAseq (p=0.86). The diagnostic pathway and final diagnostic method for the individuals in the cohort are summarized in Figure 3.

**Figure 3.**
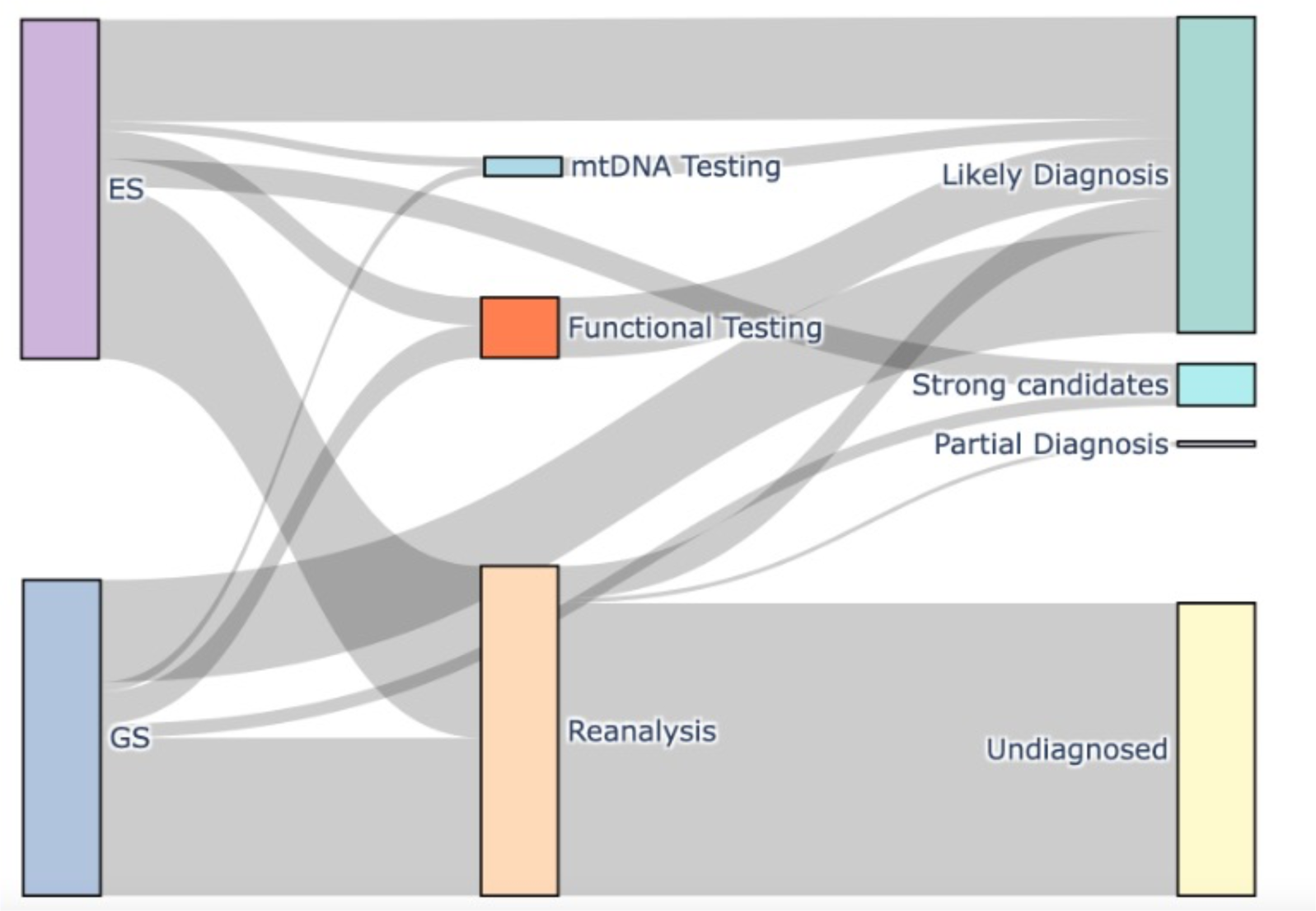
Diagnostic status by genomic testing pathway. Sankey diagram representing the diagnostic trajectory of individuals from the Mitochondrial flagship cohort. The arc’s thickness represents the proportion of individuals transitioning from analysis groups and diagnostic status.

Among the 140 patients, the adult-onset group had a 31% (n=17) molecular diagnosis rate, while the pediatric-onset group achieved a higher rate of 71% (n=60). Within the pediatric subgroups, the diagnostic yield appeared highest in the Childhood (1-5 years) group at 82%(n=14), followed by 70% (n=37) in the Infantile (<1 year) group, and 62%(n=8) in the Juvenile (5-16 years) group. The pediatric group had higher MNC scores (median 6 IQR 3) than the adult group (median 5 IQR 2) (p= 0.0005), and a higher MNC score was associated with a greater rate of molecular diagnosis in the pediatric but not in the adult participants. In addition, the MNC scores were higher in individuals with a likely diagnosis in a MD gene than in a non-mitochondrial disease gene or in the undiagnosed group (Figure 4).

**Figure 4.**
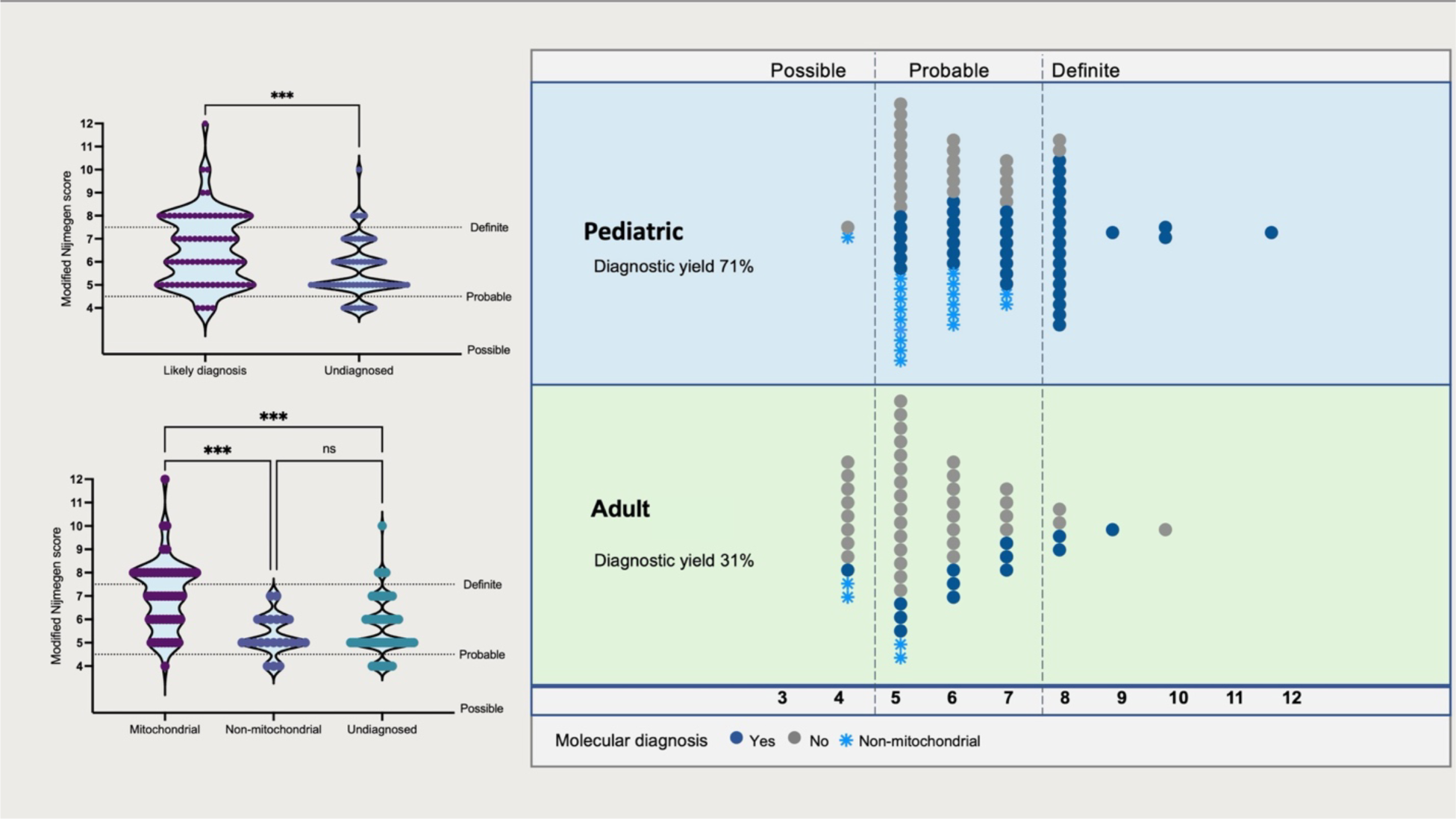
Modified Nijmegen score and diagnostic outcomes. The modified Nijmegen scores were higher in individuals with a genetic diagnosis. A higher score was associated with a mitochondrial diagnosis than a non-mitochondrial diagnosis or those who remained undiagnosed. Modified Nijmegen scores were higher in individuals with a likely molecular diagnosis in the pediatric group (median 7 IQR 3 vs 6 IQR 2 p=0.01), but not in the adult individuals (median 6 IQR 2 vs 5 IQR 1 p=0.30). *** <0.001; ns non-significant; IQR Interquartile Range.

The diagnostic yield between individuals who either reported European or non-European ancestries was similar in both age of onset groups (Table S5). Within the pediatric-onset group which had more diversity, 67% (n=26) of reported European and 78% (n=32) of non-European individuals received a molecular diagnosis. The most frequent mode of inheritance was autosomal recessive (AR), representing 42% (n=11) in European and 53% (n=17) in the non-European pediatric individuals. For the entire cohort, AR accounted for 47%, autosomal dominant (AD) for 26%, mitochondrial for 22%, dual mitochondrial and AR for 1%, and X-linked (XL) for 4%.

## D) DISCUSSION

Our results show the diagnostic utility of starting the diagnostic pathway with genomic sequencing (GS or ES+mtDNA) from blood for the diagnosis of MD. Interestingly, the diagnostic yield was higher in individuals with pediatric-onset compared to those with adult-onset (71% vs 31%, p<0.0001). Several factors likely contributed to this outcome.

Firstly, in adult blood, heteroplasmy levels for some mtDNA variants can decline with age along with mtDNA deletions becoming undetectable due to the positive selection of hematopoietic stem cells that harbor zero or a low amount of deleted mtDNA.^32-34^ The use of blood as a source of DNA testing could also be a contributing factor to why only 29% (n=5) of adult-onset individuals had a molecular diagnosis due to primary mtDNA variants in the cohort, which is lower than estimates of up to 75% of adult-onset MD being caused by mtDNA variants from previous retrospective studies.^7^ Skeletal muscle tissue was available from 12 individuals in our cohort who lacked a confirmed diagnosis after genomic testing in blood, and three of these had single mtDNA deletions detected by muscle mtDNAseq. A fourth (P47), had the m.3243A>T(*MT-TL1*) SNV detected in blood at 2% heteroplasmy, which we regarded as too low to be diagnostic but its presence at 69% heteroplasmy in muscle confirmed the genotype/phenotype relationship. Testing muscle in further individuals from the cohort could help clarify the proportion of patients in whom a diagnosis was missed due to blood-derived DNA being the initial source for testing. A recent cohort of individuals with adult-onset MD achieved a diagnostic yield of 54% (130/242).^35^ In 62% (n=80/130) of those diagnoses, the cause was mtDNA in origin. All mtDNA SNVs were detected in blood, albeit nine at heteroplasmy levels of <=3%, and seven individuals had mtDNA deletions detected in muscle that were not initially identified in blood when tested using GS.^35^

Based on these findings, we recommend muscle biopsy for patients presenting with phenotypes indicative of mtDNA deletions, such as progressive external ophthalmoplegia (PEO), or PEO plus including muscle weakness and exercise intolerance, as well as for adults with a strong clinical suspicion of mitochondrial cytopathy when blood tests are inconclusive. Muscle biopsy can uncover mtDNA deletions that may not be detectable in blood (Figure S2), as demonstrated by identifying mtDNA deletions in three individuals as well as confirming higher heteroplasmy levels in muscle tissue for one individual with low heteroplasmy levels of the m.3243A>T (*MT-TL1*) variant in blood.

A second contributor to the lower diagnostic yield in adults could be the selection of individuals, whereby adults with well-defined mitochondrial phenotypes may have already undergone targeted molecular testing rather than being recruited into this study. Targeted testing for the most common pathogenic mtDNA SNVs (e.g., m.3243A>G in *MT-TL1*, m.1555A>G in *MT-RNR1*, m.11778G>A in *MT-ND4*, m.14484T>C in *MT-ND6*, m.3460G>A in *MT-ND1*) has been available for decades and no individuals with these SNVs were detected in the Mitochondrial flagship cohort.^2^ Therefore, it is likely that individuals with pathogenic mtDNA SNVs causing common mtDNA disorders such as Mitochondrial myopathy, Encephalopathy, Lactic Acidosis, and Stroke-like episodes (MELAS, MIM 540000) or Leber Hereditary Optic Neuropathy (LHON, MIM 535000) had prior testing and were not recruited to this cohort.^2^

A third contributor to the lower diagnostic yield in adults is that the MNC score used in our inclusion criteria appears less useful for adult-onset than pediatric-onset individuals. The original Nijmegen criteria were developed as a diagnostic tool to evaluate the likelihood of a child having MD and neither the original criteria nor the MNC have been validated for adults.^14^ In a recent cohort of adult-onset MD, a higher Nijmegen criteria score was not found to be associated with a diagnosis.^35^ In the Mitochondrial flagship cohort, the MNC score was a useful tool to prioritize which pediatric-onset individuals would be more likely to receive a molecular diagnosis. However, different diagnostic and prioritization criteria may be required for adults and further research involving larger cohorts is necessary to develop appropriate screening tools for this population.

The MNC score does appear to be a useful tool for identifying individuals who have a higher likelihood of a molecular diagnosis in genes related to mitochondrial function. For the 77 individuals with a likely molecular diagnosis, the cause was in a known MD gene rather than a phenocopy gene in 100% (24/24) of individuals with a MNC score >7 (“definite”), compared to 61% (30/49) in the probable and 25% (1/4) in the possible groups.

Expanding beyond known mitochondrial disease genes in individuals with a MNC score <8 resulted in the identification of 29% (n=22) of the molecular diagnoses, which is comparable with findings from another highly selected cohort of 40 pediatric individuals with suspected mitochondrial disease, where non-mitochondrial disease genes accounted for 18% (7/40) of diagnoses.^6^ In cohorts with less stringent inclusion criteria, non-mitochondrial disorders were even more common than mitochondrial disorders (63% of diagnoses).^12^

A diagnosis in a non-mitochondrial disease gene was identified even in individuals where imaging or biochemical evidence was suggestive of a MD. Three children were diagnosed with *MORC2-*neurodevelopmental disease (MIM 619090) and one adult with Alpha-methylacyl-CoA racemase (AMACR) deficiency (MIM 614307). *De novo* variants in *MORC2* (HGNC:23573) have recently emerged as a mitochondrial phenocopy gene, with some individuals having Leigh syndrome-like lesions on brain MRI.^36^ Similarly, *AMACR* (HGNC:451) variants have also been recognized in multiple adults with suspected MD.^37^ In addition, an individual with persistent 3-methylglutaconic aciduria (3MGA), a biomarker often associated with phospholipid remodeling or mitochondrial membrane-associated disorders,^38^ was diagnosed with Kleefstra syndrome (MIM 607001), which is associated with a gene (*EHMT1;* HGNC:24650) not known to cause mitochondrial disease.^39^ Testing for 3MGA in additional patients with Kleefstra syndrome could clarify if there is an underlying secondary mechanism associated with the persistent 3MGA. Overall, these examples highlight the utility of non-targeted sequencing approaches and expanding analyses to include genes without a known mitochondrial function.

A molecular diagnosis is yet to be identified in 45% of individuals who were part of this study. This is likely due to a combination of factors: firstly, technological limitations make it difficult to identify certain types of genetic variations particularly in ES data (such as SV, short tandem repeats, and variants in non-coding regions). Secondly, many challenges are related to limitations of variant interpretation. For instance, while GS can technically identify variants in non-coding regions, our ability to accurately interpret these variants is still evolving. Bioinformatic approaches are improving rapidly; for instance, when the Mitochondrial flagship program first began, SpliceAI^40^ — a tool for analyzing and interpreting genetic variation that might affect the splicing process, was not yet available. Currently, it is considered a standard tool for variant filtration and prioritization. The re-analysis of existing genomic data, as more variant interpretation tools become available and novel disease genes are discovered, is expected to be a valuable tool for increasing diagnostic yield.^41,42^ A meta-analysis estimated a 10% increase in diagnostic yield (95% CI = 6–13%) by reanalyzing genomic data after approximately 24 months.^43^ Consistent with these findings, our reanalysis resulted in 5.7% increase in diagnostic yield. While this highlights the value of regular reanalysis, this process is mostly manual, iterative, and labor-intensive due to the complexities involved in handling large datasets and variant interpretation. Semi-automated systems are currently being developed that will streamline reanalysis efforts, likely improving both efficiency and manageability. ^44,45^

Although we conducted singleton analyses in our study, we recognize that trio analysis facilitates variant interpretation by providing segregation data and enabling the identification of *de novo* variants, which can increase the speed of diagnosis. While funding remains a primary barrier, the decreasing costs of sequencing make trio exome/genome sequencing a reliable option for first-tier diagnostic testing, particularly for patients with suspected mitochondrial disease.

Enhancing diversity in population genomic databases and addressing the overrepresentation of European ancestries in genetic studies are expected to significantly improve the variant filtration process.^46^ In the Mitochondrial flagship study, despite similar diagnostic yield between individuals with reported European and Non-European ancestries (Table S5), applying standard variant filtration criteria (AF <0.01, moderate/high impact variants, GQ > 20, AB > 0.2) resulted in a higher number of rare coding variants in non-European individuals compared to those with European ancestry, with median variant counts being 342 (IQR 103) versus 235 (IQR 41), respectively (Figure S3). This reinforces the need for inclusive genomic research efforts to expand genetic databases and better represent global population diversity.

Combining GS with other methodologies, such as transcriptome, proteome, metabolome, lipidome, and glycome analyses, may help to overcome some of the limitations of using ES/GS alone. For example, RNA sequencing (RNAseq) can detect abnormal gene expression, mono-allelic expression, or splicing defects, while quantitative proteomics can detect changes in protein abundance for different variant types, including missense, intronic and copy number variants as well as downstream effects of these variants on pathways and complexes.^29,47-50^ Similarly, studies of metabolites, lipids, and glycans can detect characteristic metabolite profiles and biomarkers.^51^

Consequently, individuals with high MNC scores who are still molecularly undiagnosed are currently being recruited to other research projects to incorporate systematic reanalysis and other -omic technologies with the aim to provide more patients and families with a molecular diagnosis.

Building on the effort to extend molecular diagnoses to more individuals, the Mitochondrial flagship has contributed to shaping standard care in Australia. By providing data to Australia’s Medical Services Advisory Committee (MSAC) recommendation application 1675,^52^ it has played a role in the establishment of new Medicare Benefits Schedule (MBS) item numbers (73456, 73457, 73458, 73459, 73460, 73461 and 73462)^53^ which support genomic testing for suspected mitochondrial disease as part of Australia’s universal health insurance scheme. This initiative is a major step forward in promoting diagnostic accuracy and equal access to genomic testing through public funding.

## Data Availability

The datasets supporting the current study have not been deposited in a public repository due to consent restrictions. De-identified genomic and associated data from this study are available for ethically approved research. The online access application process is administered by the Australian Genomics Data Access Committee. For queries about the data sets please contact. All class 4 and 5 variants described here were deposited in ClinVar before publication.

## Acknowledgments

We would like to thank all the participants and families who participated in this study, and the clinical teams involved in their care. The research conducted at the Murdoch Children’s Research Institute was supported by the Victorian Government’s Operational Infrastructure Support Program. The Chair in Genomic Medicine awarded to JC is generously supported by The Royal Children’s Hospital Foundation. We acknowledge the Bio21 Mass Spectrometry and Proteomics Facility (MMSPF) for the provision of instrumentation, training, and technical support.

## Author Contributions

Conceptualization- J.C., D.R.T

Data curation- R.R., A.G.C., N.L.B., L.E.F., N.J.L., J.E.M.

Formal analysis- R.R., A.G.C., L.E.F., A.E.F., A.E.W., D.H.H., D.A.S., I.G, Y.W.

Funding acquisition- J.C., D.R.T., T.F.B.

Investigation-S.Be, J.Br., J.C.L., K.Bo., J.Bu., A.B., S.E., M.G.dS, K.F., D.H.H., D.H., M.R.J.,

M.K., S.K-S., M.M., E.M.M., D.A.S., J.L.,M.C.J.Q., S.S., T.F.B., S.C., M.H., L.E.F., A.E.W., A.G.C., R.R., A.E.F.

Methodology- M.J.C, C.S., M.T.R.

Project administration- N.L.B., M.G.dS, T.F.B., T.M., S.M.,

Clinical intake committee- S.Ba., D.B., K.Bh., D.C., J.C., M.B.D., C.E., J.F., M.C.F., R.G., H.G.,

M.P.K., P.J.L., J.L., J.McG., J.P., L.K.P., N.S., D.R.T., M.C.T., M.W., T.L.W., C.W.

Resources- J.C., D.R.T, C.E

Software- M.J.B., A.M-J., M.J.C, C.S., V.G.

Supervision- J.C., D.R.T, A.G.C.

Validation- B.C., S.C.

Visualization- R.R.

Writing-original draft- R.R.

Writing-review & editing- J.C., A.G.C., D.R.T. and all authors

## Ethics Declaration

This study was conducted in accordance with the revised Declaration of Helsinki and following the Australian National Health and Medical Research Council statement of ethical conduct in research involving humans. The Mitochondrial flagship study was reviewed and approved through our lead Human Research Ethics Committee (HREC), Royal Melbourne Hospital (formerly known as Melbourne Health) (HREC/16/MH/251). Sites that were not covered at the time by the Australian National Mutual Acceptance system were reviewed and approved by the Western Australian Child and Adolescent Health Service HREC (RGS0000000086), Tasmanian HREC (H0016443) and Queensland UnitingCare Health HREC (1717).

