## Supplementary Figures for "The Australian Genomics Mitochondrial Flagship: A National Program Delivering Mitochondrial Diagnoses"

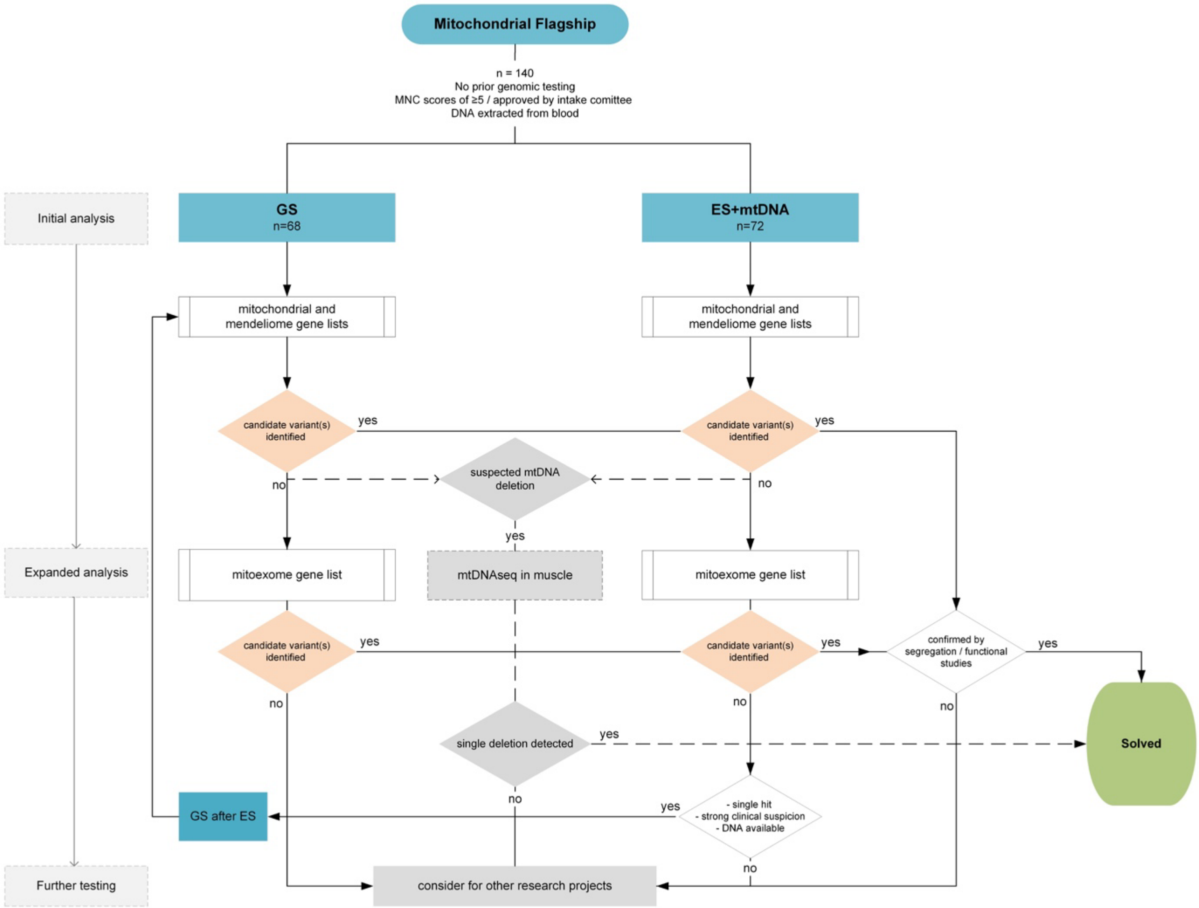


**Figure S1. Mitochondrial Flagship Flowchart**

Variants were prioritized using PanelApp Australia's mitochondrial disease and mendeliome gene lists. Unresolved individuals underwent expanded analysis with a custom mitoexome list, including genes with evidence of mitochondrial function but not yet linked to monogenic diseases. Muscle mtDNA testing was recommended if the phenotype was compatible with a mtDNA deletion or a low heteroplasmy variant in blood. Individuals who remain molecularly undiagnosed are being enrolled in other research projects for systematic reanalysis and the application of additional -omic technologies.

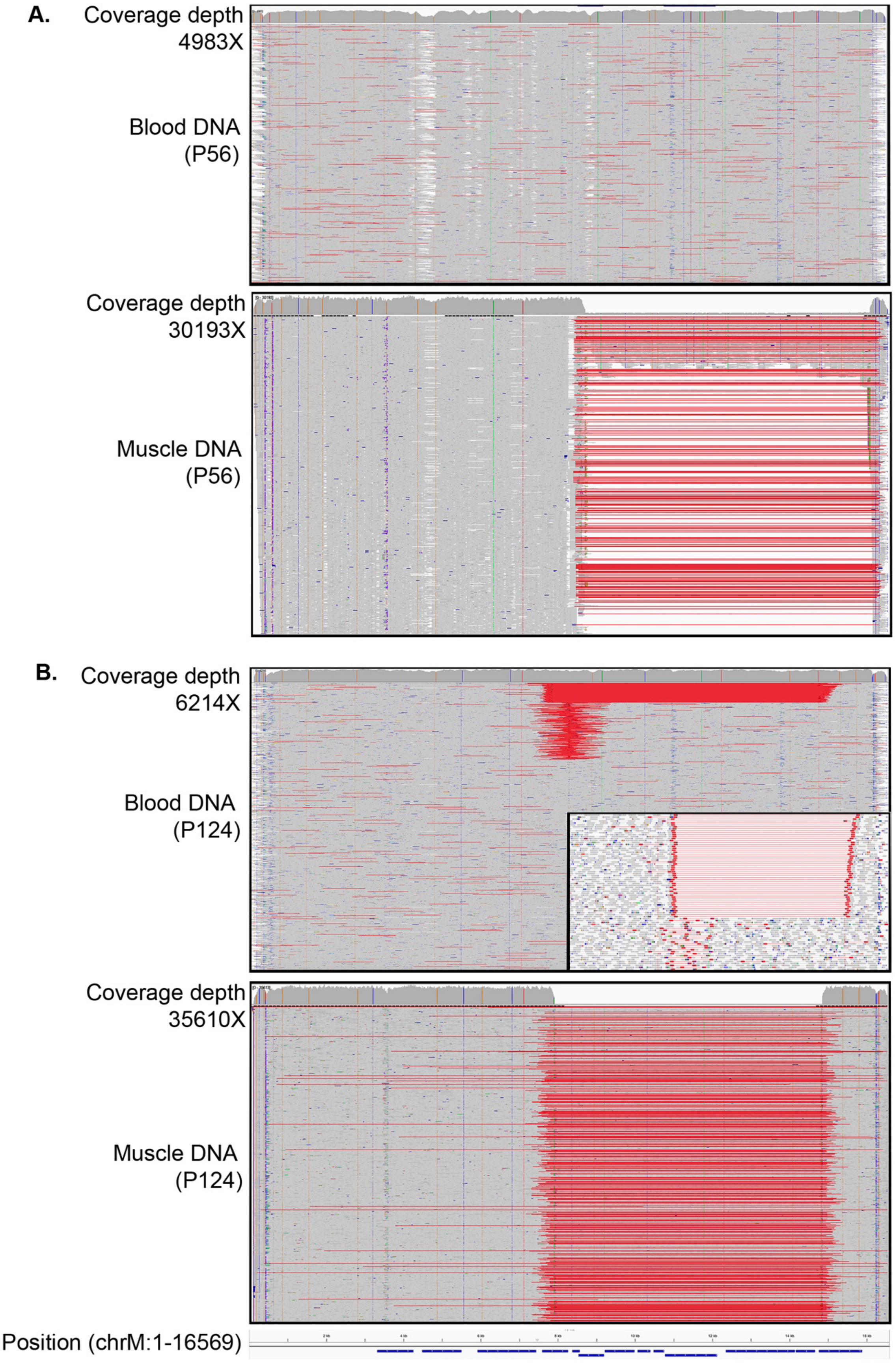

A.
Coverage depth
4983X
Blood DNA
(P56)
Coverage depth
30193X
Muscle DNA
(P56)
B.
Coverage depth
6214X
Blood DNA
(P124)
Coverage depth
35610X
Muscle DNA
(P124)
Position (chrM:1-16569)

**Figure S2. Pathogenic mtDNA deletions were identified in DNA extracted from skeletal muscle but undetectable or below reportable diagnostic thresholds in blood DNA.**

IGV coverage plots showing the mtDNA sequencing derived from blood and muscle for P56 (A) and P124 (B) with coverage depths indicated. (A) The pathogenic mtDNA deletion spanning 7.44kb (chrM:8649-16084) was not detected in the blood DNA for P56 but was in 80-90% of the reads in the muscle DNA (as indicated by the red lines). (B) The pathogenic mtDNA deletion spanning 6.98kb (chrM:7821-14798) was initially not identified in the blood DNA for P124 however data were reanalysed after the deletion was detected at 80-90% heteroplasmy in skeletal muscle. Reanalysis showed that the deletion was present in blood in 53 reads of 6214 (inset shows this in detail) or ~0.8% heteroplasmy, below reportable levels.

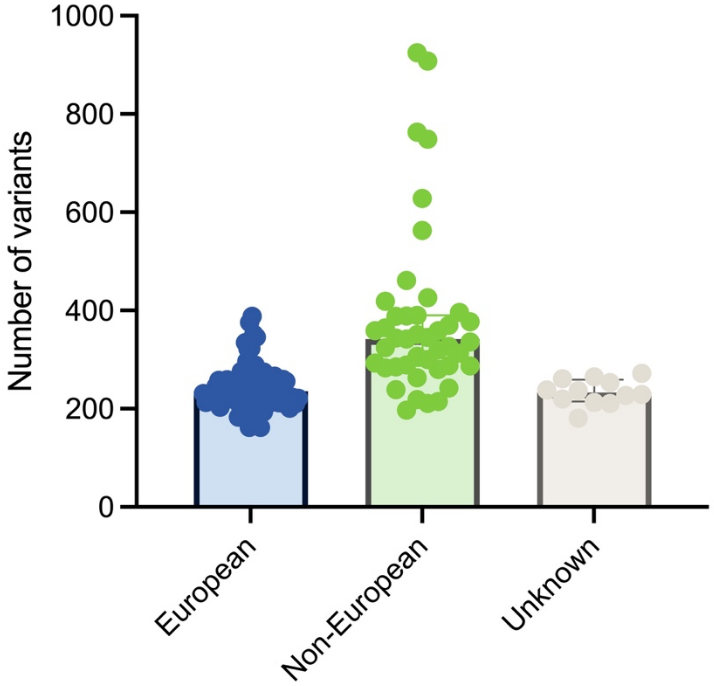


**Figure S3. Number of rare variants in individuals with Non-European and European reported ancestry.**

Using variant filtration criteria that included allele frequency <0.01, moderate/high impact variants, Genotype quality > 20, allele balance > 0.2, resulted in a higher number of rare coding variants in individuals with reported non-European ancestry (median 342, IQR 103) compared to those with reported European ancestry (median 235, IQR 41).
