## Supplementary material for "The Australian Genomics Mitochondrial Flagship: A National Program Delivering Mitochondrial Diagnoses": Table S1

Table S1 Modified Nijmegen criteria

| I. Clinical criteria (max. 4 points) | | | II. Metabolic and imaging studies  (max. 4 points) | III. Morphology and biochemical  (max. 4 points) |
| --- | --- | --- | --- | --- |
| A. Muscular presentation  (max. 2 points) | **B. CNS presentation**  **(max. 2 points)** | **C. Multisystem disease**  **(max. 3 points)** |  |  |
| - Progressive external ophthalmoplegia *^a^* - Facies myopathica - Ptosis^*^ - Exercise intolerance - Muscle weakness - Rhabdomyolysis - Abnormal EMG | - Intellectual disability^*^ - Loss of skills / regression - Stroke-like episode - Migraine - Seizures - Myoclonus - Cortical blindness - Optic neuropathy^*^ - Retinitis pigmentosa^*^ - Pyramidal signs - Extrapyramidal signs - Brainstem involvement | - Haematology - GI tract - Endocrine / growth - Diabetes mellitus^*^ - Cardiomyopathy - Hypertension^*^ - Kidney - Sensorineural deafness^*^ - Neuropathy - Recurrent /familial | - Elevated blood lactate *^b*^* - Elevated L/P ratio - Elevated blood alanine - Elevated serum FGF21*^a*^* - Elevated serum GDF15*^a*^* - Elevated CSF lactate - Elevated CSF protein - Elevated CSF alanine - Urinary TA excretion - Ethylmalonic aciduria - 2-ethylhydracylic aciduria^*^ - 3-methylglutaconic aciduria^*^ - Stroke-like picture/MRI *^a^* - Leigh syndrome/MRI *^a^* - Elevated lactate/MRS *^a^* | - Ragged red/blue fibres *^c^* - COX-negative fibres - Reduced COX staining - SDH positive blood vessels *^a^* - Abnormal mitochondria/EM *^a^* - Abnormal RC enzymology *^d*^* - mtDNA depletion or multiple mtDNA deletions *^a*^* |

*^a^* This specific symptom scores 2 points.

*^b^* If blood lactate is elevated once scores 1 point; if elevated thrice scores 2 points.

*^c^* 2 points if present; 4 points if >2%.

*^d^* Score 2 points if <20% residual activity (relative to marker enzymes such as citrate synthase or RC complex II) of any RC complex in a tissue or <30% residual activity of any RC complex in a cell line or <30% residual activity of any RC complex in two or more tissues. Score 1 point if 20–30% residual activity of any RC complex in a tissue or 30–40% residual activity of any RC complex in a cell line or 30–40% residual activity of any RC complex in two or more tissues.

* Modified criteria based on Morava et al., 2006 and Riley et al., 2020.
