## Supplementary material for "The Australian Genomics Mitochondrial Flagship: A National Program Delivering Mitochondrial Diagnoses": Table S3

| n | Individual | Sex | Initial analysis | Age at Onset | MNC | Likely Diagnosis | Molecular Diagnosis | Diagnostic testing pathway | Gene | HGNC ID | Variants (Zygosity / heteroplasmy (%) / classification ) | Mode of Inheritance | Diagnosis in mitochondrial disease gene | HPO terms ( top 5 *) | Functional studies | Functional studies - type | Previously published |
|---|---|---|---|---|---|---|---|---|---|---|---|---|---|---|---|---|---|
| 1 | P1 | Male | GS | pediatric | 8 | Yes | positive | solved by initial analysis (GS/ESmtDNAseq incl. segregation) | *TPK1* | HGNC:17358 | NM_022445.4:c.479C>A p.(Ser160Ter) het 5<br>NM_022445.4:c.161A>G p.(Asp54Gly) het 5 | AR | Yes | HP:0003324 Generalized muscle weakness; HP:0001332 Dystonia; HP:0002376 Developmental regression; HP:0002500 Abnormal cerebral white matter morphology ; HP:0012751 Abnormal basal ganglia MRI signal intensity | No | | |
| 2 | P2 | Female | GS | adult | 8 | Yes | positive | solved by initial analysis (GS/ESmtDNAseq incl. segregation) | *OPA1* | HGNC:8140 | NM_130837.3:c.1499G>A p.(Arg500His) het 5 | AD | Yes | HP:0000508 Ptosis; HP:0000602 Ophthalmoplegia; HP:0003324 Generalized muscle weakness; HP:0000407 Sensorineural hearing impairment ; HP:0000648 Optic atrophy | No | | |
| 3 | P3 | Female | GS | pediatric | 5 | Yes | positive | solved by functional testing | *SERAC1* | HGNC:21061 | NC_000006.11:g.158547971_158556962del<br>NP_116250.3:p.(Gly247AlafsTer10) het 4<br>NM_032861.4:c.548G>A p.(Arg183Gln) het 4 | AR | Yes | HP:0001260 Dysarthria; HP:0001332 Dystonia; HP:0002376 Developmental regression; HP:0001399 Hepatic failure; HP:0012751 Abnormal basal ganglia MRI signal intensity | Yes | cDNA studies (liver)<br>Cloning experiments | |
| 4 | P4 | Male | ES | pediatric | 5 | No | undiagnosed | unsolved | | | | | | HP:0001332 Dystonia; HP:0001249 Intellectual disability ; HP:0001250 Seizure; HP:0000407 Sensorineural hearing impairment ; HP:0012751 Abnormal basal ganglia MRI signal intensity | | | |
| 5 | P5 | Male | ES | pediatric | 6 | Yes | positive | solved by functional testing | *MOGS, CRYAA* | HGNC:24862,<br>HGNC:2388 | NM_006302.3:c.1225G>A p.(Gly409Arg) hom 4<br>NM_000394.4:c.346C>T p.(Arg116Cys) het 5 | AR | No | HP:0001257 Spasticity; HP:0001263 Global developmental delay; HP:0001250 Seizure; HP:0000407 Sensorineural hearing impairment ; HP:0000518 Cataracts; | Yes | glycomics | PMID: 36651519 |
| 6 | P6 | Female | GS | adult | 5 | No | undiagnosed | unsolved | | | | | | HP:0001324 Muscle weakness; HP:0003546 Exercise intolerance; HP:0003236 Elevated creatine kinase ; HP:0002151 Increased serum lactate; HP:0003688 Cytochrome C oxidase-negative muscle fibers | | | |
| 7 | P7 | Female | ES | pediatric | 5 | Yes | positive | solved by initial analysis (GS/ESmtDNAseq incl. segregation) | *SLC2A1* | HGNC:11005 | NM_006516.4:c.481C>T p.(Gln161Ter) het 5 | AD | No | HP:0000508 Ptosis; HP:0001251 Ataxia; HP:0002376 Developmental regression; HP:0001250 Seizure; HP:0002500 Abnormal cerebral white matter morphology | No | | |
| 8 | P8 | Male | ES | adult | 5 | Yes | positive | solved by muscle mtDNA testing | *mtDNA deletion* | | mtDNA deletion (7-8kb detected by Southern Blot) hetp (% not quantified) 5 | Mitochondrial | Yes | HP:0000602 Ophthalmoplegia; HP:0002076 Migraine ; HP:0009830 Peripheral neuropathy; HP:0000648 Optic atrophy; HP:0001638 Cardiomyopathy; | No | | |
| 9 | P9 | Female | GS | pediatric | 8 | No | undiagnosed | unsolved | | | | | | HP:0003324 Generalized muscle weakness; HP:0001332 Dystonia; HP:0002376 Developmental regression; HP:0001250 Seizure; HP:0002500 Abnormal cerebral white matter morphology; | | | |
| 10 | P10 | Female | ES | adult | 5 | No | undiagnosed | unsolved | | | | | | HP:0000508 Ptosis; HP:0000407 Sensorineural hearing impairment ; HP:0000510 Retinitis pigmentosa; HP:0001733 Pancreatitis ; HP:0030890 White matter hyperintensities; | | | |
| 11 | P11 | Female | ES | pediatric | 8 | Yes | positive | solved by initial analysis (GS/ESmtDNAseq incl. segregation) | *MT-ND5* | HGNC:7461 | NC_012920.1:m.13513G>A p.(Asp393Asn) hetp (64%) 5 | Mitochondrial | Yes | HP:0000508 Ptosis; HP:0001283 Bulbar palsy; HP:0001263 Global developmental delay; HP:0002376 Developmental regression; HP:0012751 Abnormal basal ganglia MRI signal intensity; | No | | |
| 12 | P12 | Male | ES | adult | 8 | No | undiagnosed | unsolved | | | | | | HP:0000508 Ptosis; HP:0003326 Myalgia; HP:0002015 Dysphagia; HP:0000407 Sensorineural hearing impairment ; HP:0100704 Cerebral visual impairment ; | | | |
| 13 | P13 | Female | ES | pediatric | 8 | Yes | positive | solved by initial analysis (GS/ESmtDNAseq incl. segregation) | *TSFM* | HGNC:12367 | NM_005726.6:c.774G>C p.(Gln258His) het 4<br>NM_005726.6:c.856C>T p.(Gln286Ter) het 5 | AR | Yes | HP:0001263 Global developmental delay; HP:0000407 Sensorineural hearing impairment ; HP:0001638 Cardiomyopathy; HP:0001508 Failure to thrive ; HP:0004322 Short stature ; | No | | |
| 14 | P14 | Female | GS | pediatric | 5 | Yes | positive | solved by initial analysis (GS/ESmtDNAseq incl. segregation) | *MT-ND1* | HGNC:7455 | NC_012920.1:m.3697G>A p.(Gly131Ser) hetp (69%) 5 | Mitochondrial | Yes | HP:0003324 Generalized muscle weakness; HP:0002015 Dysphagia; HP:0001332 Dystonia; HP:0002376 Developmental regression; HP:0012751 Abnormal basal ganglia MRI signal intensity; | No | | |
| 15 | P15 | Female | GS | pediatric | 5 | No | undiagnosed | unsolved | | | | | | HP:0003546 Exercise intolerance; HP:0001252 Hypotonia ; HP:0001263 Global developmental delay; HP:0001249 Intellectual disability; HP:0008322 Abnormal mitochondrial morphology; | | | |
| 16 | P16 | Male | ES | pediatric | 5 | Yes | positive | solved by functional testing | *ATAD3A*<br>*ATAD3B*<br>*ATAD3C* | HGNC:25567<br>HGNC:24007<br>HGNC:32151 | NC_000001.11:g.1456616-1524663dup het 5 | AD | Yes | HP:0001639 Hypertrophic cardiomyopathy ; HP:0001706 Endocardial fibroelastosis; HP:0002151 Increased serum lactate; HP:0003535 3-methylglutaconic aciduria; HP:0012751 Abnormal basal ganglia MRI signal intensity; | Yes | Proteomics, cDNA studies, qRT-PCR | PMID: 33575671 |
| 17 | P17 | Male | ES | adult | 4 | Yes | positive | solved by initial analysis (GS/ESmtDNAseq incl. segregation) | *RRM2B* | HGNC:17296 | NM_015713.5:c.979C>T p.(Arg327Ter) het 5 | AD | Yes | HP:0000508 Ptosis; HP:0000602 Ophthalmoplegia; HP:0001324 Muscle weakness; HP:0003546 Exercise intolerance; HP:0000407 Sensorineural hearing impairment; | No | | |
| 18 | P18 | Male | ES | pediatric | 8 | Yes | positive | solved by initial analysis (GS/ESmtDNAseq incl. segregation) | *MT-TV* | HGNC:7500 | NC_012920.1:m.1638T>C homp 4 | Mitochondrial | Yes | HP:0001639 Hypertrophic cardiomyopathy ; HP:0000122 Unilateral renal agenesis; HP:0001903 Anaemia; HP:0002151 Increased serum lactate; HP:0003348 Hyperalaninemia; | No | | |
| 19 | P19 | Female | ES | adult | 4 | No | undiagnosed | unsolved | | | | | | HP:0001332 Dystonia; HP:0001251 Ataxia; HP:0000648 Optic atrophy; HP:0000819 Diabetes mellitus; HP:0002401 Stroke-like episode | | | |
| 20 | P20 | Female | GS | adult | 6 | No | undiagnosed | unsolved | | | | | | HP:0030880 Raynaud phenomenon; HP:0000508 Ptosis; HP:0001324 Muscle weakness; HP:0003324 Generalized muscle weakness; HP:0009830 Peripheral neuropathy; | | | |
| 21 | P21 | Female | ES | pediatric | 8 | Yes | positive | solved by initial analysis (GS/ESmtDNAseq incl. segregation) | *COX15* | HGNC:2263 | NM_078470.6:c.839T>C p.(Phe280Ser) het 4<br>NM_078470.6:c.287G>T p.(Gly96Val) het 4 | AR | Yes | HP:0000508 Ptosis; HP:0002015 Dysphagia; HP:0001263 Global developmental delay; HP:0001250 Seizure; HP:0003535 3-methylglutaconic aciduria; | No | | |
| 22 | P22 | Female | GS | adult | 4 | No | undiagnosed | unsolved | | | | | | HP:0001324 Muscle weakness; HP:0003546 Exercise intolerance; HP:0002076 Migraine ; HP:0000407 Sensorineural hearing impairment ; HP:0002500 Abnormal cerebral white matter morphology | | | |
| 23 | P23 | Female | ES | adult | 5 | Yes | positive | solved by initial analysis (GS/ESmtDNAseq incl. segregation) | *AMACR* | HGNC:451 | NM_014324.6:c.154T>C p.(Ser52Pro) hom 5 | AR | No | HP:0002076 Migraine ; HP:0001250 Seizure; HP:0100704 Cerebral visual impairment ; HP:0000510 Retinitis pigmentosa; HP:0012751 Abnormal basal ganglia MRI signal intensity | No ++ | | |
| 24 | P24 | Female | GS | pediatric | 4 | No | undiagnosed | unsolved | | | | | | HP:0001251 Ataxia; HP:0001250 Seizure; HP:0000407 Sensorineural hearing impairment ; HP:0000648 Optic atrophy; HP:0004322 Short stature | | | |
| 25 | P25 | Female | ES | pediatric | 8 | Yes | positive | solved by initial analysis (GS/ESmtDNAseq incl. segregation) | *MT-TW* | HGNC:7501 | NC_012920.1:m.5559A>G homp 5 | Mitochondrial | Yes | HP:0003324 Generalized muscle weakness; HP:0001257 Spasticity; HP:0002401 Stroke-like episode; HP:0001250 Seizure; HP:0001399 Hepatic failure; | No | | |
| 26 | P26 | Male | GS | pediatric | 7 | No | undiagnosed | unsolved | | | | | | HP:0003324 Generalized muscle weakness; HP:0001638 Cardiomyopathy; HP:0002151 Increased serum lactate; HP:0008322 Abnormal mitochondrial morphology ; HP:0008972 Decreased activity of mitochondrial respiratory chain | | | |
| 27 | P27 | Male | ES | pediatric | 5 | No | undiagnosed | unsolved | | | | | | HP:0003324 Generalized muscle weakness; HP:0001249 Intellectual disability ; HP:0002151 Increased serum lactate; HP:0012747Abnormal brainstem MRI signal intensity | | | |
| 28 | P28 | Female | ES | adult | 6 | No | undiagnosed | unsolved | | | | | | HP:0003324 Generalized muscle weakness; HP:0003546 Exercise intolerance; HP:0003236 Elevated creatine kinase ; HP:0002076 Migraine ; HP:0000822 Hypertension; | | | |
| 29 | P29 | Male | ES | pediatric | 6 | No | undiagnosed | unsolved | | | | | | HP:0001263 Global developmental delay; HP:0001250 Seizure; HP:0000510 Retinitis pigmentosa; HP:0001903 Anaemia; HP:0002453Abnormal globus pallidus morphology; | | | |
| 30 | P30 | Male | ES | adult | 6 | Yes | positive | solved by initial analysis (GS/ESmtDNAseq incl. segregation) | *SPG7* | HGNC:11237 | NM_003119.4:c.1529C>T p.(Ala510Val) hom 5 | AR | Yes | HP:0000602 Ophthalmoplegia; HP:0000510 Retinitis pigmentosa; HP:0003200Ragged-red muscle fibers; HP:0003688 Cytochrome C oxidase-negative muscle fibers | No | | |
| 31 | P31 | Female | GS | pediatric | 5 | Yes | positive | solved by initial analysis (GS/ESmtDNAseq incl. segregation) | *DHCR7* | HGNC:2860 | NM_001360.3:c.964-1G>C het 5<br>NM_001360.3:c.866C>T p.(Thr289Ile) het 5 | AR | No | HP:0001263 Global developmental delay; HP:0001250 Seizure; HP:0000407 Sensorineural hearing impairment ; HP:0010569 Elevated 7-dehydrocholesterol ; HP:0000252 Microcephaly | No + | | |
| 32 | P32 | Male | GS | pediatric | 8 | Yes | positive | solved by initial analysis (GS/ESmtDNAseq incl. segregation) | *PDHA1* | HGNC:8806 | NM_000284.4:c.772G>C p.(Asp258His) hem 4 | XL | Yes | HP:0000508 Ptosis; HP:0001251 Ataxia; HP:0001250 Seizure; HP:0002151 Increased serum lactate; HP:0012751 Abnormal basal ganglia MRI signal intensity; | No | | |
| 33 | P33 | Male | ES | adult | 6 | Yes | positive | solved by initial analysis (GS/ESmtDNAseq incl. segregation) | *MT-ND3* | HGNC:7458 | NC_012920.1:m.10191T>C p.(Ser45Pro) hetp (22%) 5 | Mitochondrial | Yes | HP:0000602 Ophthalmoplegia; HP:0001251 Ataxia; HP:0001250 Seizure; HP:0002151 Increased serum lactate ; HP:0012751 Abnormal basal ganglia MRI signal intensity | No | | |
| 34 | P34 | Female | GS | adult | 8 | Yes | positive | solved by reanalysis of GS or ES | *C1QBP* | HGNC:2494 | NM_001212.4:c.612C>G p.(Phe204Leu) hom 5 | AR | Yes | HP:0000508 Ptosis; HP:0000602 Ophthalmoplegia; HP:0000407 Sensorineural hearing impairment ; HP:0001638 Cardiomyopathy; HP:0001639 Hypertrophic cardiomyopathy; | No | | |
| 35 | P35 | Male | ES | adult | 7 | Yes | positive | solved by initial analysis (GS/ESmtDNAseq incl. segregation) | *TWNK* | HGNC:1160 | NM_021830.5:c.1374G>T p.(Gln458His) het 5 | AD | Yes | HP:0000508 Ptosis; HP:0000602 Ophthalmoplegia; HP:0003236 Elevated creatine kinase; HP:0000819 Diabetes mellitus; HP:0002151 Increased serum lactate; | No | | |
| 36 | P36 | Male | ES | adult | 6 | No | undiagnosed | unsolved | | | | | | HP:0030319 Weakness of facial musculature; HP:0001324 Muscle weakness; HP:0003546 Exercise intolerance; HP:0003236 Elevated creatine kinase ; HP:0002015 Dysphagia; HP:0001251 Ataxia; HP:0000407 Sensorineural hearing impairment ; HP:0000821 Hypothyroidism; HP:0001272 Cerebellar atrophy | | | |
| 37 | P37 | Male | ES | pediatric | 7 | Yes | positive | solved by reanalysis of GS or ES | *MORC2* | HGNC:23573 | NM_001303256.3:c.259T>C p.(Ser87Pro) het 5 | AD | No | HP:0001252 Hypotonia ; HP:0001263 Global developmental delay; HP:0009830 Peripheral neuropathy; HP:0002151 Increased serum lactate; HP:0012751 Abnormal basal ganglia MRI signal intensity; | No | | |
| 38 | P38 | Female | GS | adult | 5 | No | undiagnosed | unsolved | | | | | | HP:0000508 Ptosis; HP:0000602 Ophthalmoplegia; HP:0000819 Diabetes mellitus; HP:0000821 Hypothyroidism; HP:0000846 Adrenal insufficiency; | | | |
| 39 | P39 | Female | ES | adult | 4 | Yes | positive | solved by initial analysis (GS/ESmtDNAseq incl. segregation) | *MPZ* | HGNC:7225 | NM_000530.8:c.233C>A p.(Ser78Ter) het 5 | AD | No | HP:0001251 Ataxia; HP:0009830 Peripheral neuropathy; HP:0000407 Sensorineural hearing impairment ; HP:0031546 Cardiac conduction abnormality ; HP:0000819 Diabetes mellitus; | No | | |
| 40 | P40 | Female | GS | pediatric | 5 | Yes | positive | solved by functional testing | *PEX6* | HGNC:8859 | NM_000287.4:c.2356C>T p.(Arg786Trp) het 5<br>NM_000287.4:c.371T>C p.(Leu124Pro) het 4 | AR | No | HP:0009830 Peripheral neuropathy; HP:0000407 Sensorineural hearing impairment ; HP:0000510 Retinitis pigmentosa; HP:0000135 Hypogonadism; HP:0002500 Abnormal cerebral white matter morphology; | Yes | Allele-specific PCR , analysis of very-long-chain fatty acids (VLCFA) in plasma | PMID: 32399598 |
| 41 | P41 | Female | GS | pediatric | 6 | No | undiagnosed | unsolved | | | | | | HP:0004389 Intestinal pseudo-obstruction ; HP:0002151 Increased serum lactate; HP:0003348 Hyperalaninemia; HP:0002490 Increased CSF lactate ; HP:0025045 Abnormal brain lactate level by MRS | | | |
| 42 | P42 | Female | ES | pediatric | 7 | Yes | positive | solved by initial analysis (GS/ESmtDNAseq incl. segregation) | *NARS2* | HGNC:26274 | NM_024678.6:c.1253G>A p.(Arg418His) hom 4 | AR | Yes | HP:0001332 Dystonia; HP:0001263 Global developmental delay; HP:0001250 Seizure; HP:0000407 Sensorineural hearing impairment ; HP:0012751 Abnormal basal ganglia MRI signal intensity | No | | |
| 43 | P43 | Female | ES | pediatric | 6 | Yes | positive | solved by initial analysis (GS/ESmtDNAseq incl. segregation) | *MT-ND1* | HGNC:7455 | NC_012920.1:m.3697G>A p.(Gly131Ser) hetp (53%) 5 | Mitochondrial | Yes | HP:0001300 Parkinsonism ; HP:0002376 Developmental regression; HP:0001250 Seizure; HP:0002151 Increased serum lactate; HP:0002500 Abnormal cerebral white matter morphology; | No | | |
| 44 | P44 | Female | ES | adult | 5 | No | undiagnosed | unsolved | | | | | | HP:0001324 Muscle weakness; HP:0003546 Exercise intolerance; HP:0003236 Elevated creatine kinase ; HP:0009830 Peripheral neuropathy; HP:0003688 Cytochrome C oxidase-negative muscle fibers | | | |
| 45 | P45 | Female | GS | adult | 6 | No | undiagnosed | unsolved | | | | | | HP:0001250 Seizure; HP:0000407 Sensorineural hearing impairment ; HP:0000648 Optic atrophy ; HP:0000510 Retinitis pigmentosa; HP:0000843 Hyperparathyroidism; | | | |
| 46 | P46 | Male | ES | adult | 6 | No | undiagnosed | unsolved | | | | | | HP:0003546 Exercise intolerance; HP:0003200Ragged-red muscle fibers; HP:0003688 Cytochrome C oxidase-negative muscle fibers; HP:0008322 Abnormal mitochondrial morphology | | | |

| | | | | | | | | | | | | | | | | | |
|---|---|---|---|---|---|---|---|---|---|---|---|---|---|---|---|---|---|
| 47 | P47 | Male | GS | adult | 5 | Yes | positive | solved by muscle mtDNA testing | *SORD, MT-TL1* | HGNC:11184,<br>HGNC:7490 | NM_003104.6:c.757del p.(Ala253fs) hom 5<br>NC_012920.1:m.3243A>T (hetp blood 2% muscle 69%) 5 | Mitochondrial and AR | Yes | HP:0001250 Seizure; HP:0009830 Peripheral neuropathy; HP:0000407 Sensorineural hearing impairment ; HP:0000518 Cataracts; HP:0030890 White matter hyperintensities; | No | | PMID: 33924034 |
| 48 | P48 | Female | GS | pediatric | 5 | No | undiagnosed | unsolved | | | | | | HP:0001324 Muscle weakness; HP:0001263 Global developmental delay; HP:0002401 Stroke-like episode ; HP:0001250 Seizure; HP:0001903 Anaemia; | | | |
| 49 | P49 | Male | ES | pediatric | 8 | Yes | positive | solved by initial analysis (GS/ESmtDNAseq incl. segregation) | *ECHS1* | HGNC:3151 | NM_004092.4:c.830C>T p.(Thr277Ile) het 4<br>NM_004092.4:c.817A>G p.(Lys273Glu) het 5 | AR | Yes | HP:0001252 Hypotonia ; HP:0001332 Dystonia; HP:0001263 Global developmental delay; HP:0002151 Increased serum lactate; HP:0012751 Abnormal basal ganglia MRI signal intensity ; | No | | |
| 50 | P50 | Female | ES | pediatric | 12 | Yes | strong candidate | strong candidate - work in progress | *TOP3A* | HGNC:11992 | NM_004618.5:c.240_240+2dup hom | AR | Yes | HP:0000602 Ophthalmoplegia; HP:0003324 Generalized muscle weakness;HP:0001263 Global developmental delay ; HP:0001250 Seizure; HP:0000407 Sensorineural hearing impairment ; | Pending | cell based mitochondrial functional studies and sister chromatid exchange studies | |
| 51 | P51 | Female | GS | pediatric | 5 | No | undiagnosed | unsolved | | | | | | HP:0002240 Hepatomegaly; HP:0001903 Anaemia; HP:0002151 Increased serum lactate; HP:0003219 Ethylmalonic aciduria; HP:0003535 3-methylglutaconic aciduria; | | | |
| 52 | P52 | Female | ES | adult | 5 | Yes | positive | solved by initial analysis (GS/ESmtDNAseq incl. segregation) | *EIF2B3* | HGNC:3259 | NM_020365.5:c.260C>T p.(Ala87Val) hom 5 | AR | No | HP:0001252 Hypotonia ; HP:0001251 Ataxia; HP:0001249 Intellectual disability ;HP:0002500 Abnormal cerebral white matter morphology ; HP:0030890 White matter hyperintensities; | No | | |
| 53 | P53 | Female | GS | pediatric | 5 | Yes | positive | solved by initial analysis (GS/ESmtDNAseq incl. segregation) | *EHMT1* | HGNC:24650 | NM_024757.5:c.3519C>G p.(Tyr1173Ter) het 5 | AD | No | HP:0001252 Hypotonia ;HP:0001250 Seizure; HP:0031218 SIADH; HP:0001513 Obesity ; HP:0003535 3-methylglutaconic aciduria | No | | |
| 54 | P54 | Male | GS | pediatric | 5 | No | undiagnosed | unsolved | | | | | | HP:0000508 Ptosis; HP:0001324 Muscle weakness; HP:0001249 Intellectual disability ; HP:0004322 Short stature ; HP:0002151 Increased serum lactate; | | | |
| 55 | P55 | Male | ES | pediatric | 5 | No | undiagnosed | unsolved | | | | | | HP:0000602 Ophthalmoplegia; HP:0003236 Elevated creatine kinase ; HP:0001336 Myoclonus; HP:0001300 Parkinsonism ; HP:0001249 Intellectual disability ; | | | |
| 56 | P56 | Male | GS | adult | 7 | Yes | positive | solved by muscle mtDNA testing | *mtDNA deletion* | | NC_012920.1:m.8649_16084del hetp (% not quantified) 5 | Mitochondrial | Yes | HP:0000508 Ptosis; HP:0000602 Ophthalmoplegia; HP:0000407 Sensorineural hearing impairment ; HP:0007957 Corneal opacity; HP:0000518 Cataracts; | No | | |
| 57 | P57 | Male | ES | pediatric | 6 | No | undiagnosed | unsolved | | | | | | HP:0000508 Ptosis; HP:0003324 Generalized muscle weakness; HP:0001283 Bulbar palsy; HP:0001249 Intellectual disability ; HP:0000407 Sensorineural hearing impairment; | | | |
| 58 | P58 | Female | GS | adult | 5 | Yes | positive | solved by initial analysis (GS/ESmtDNAseq incl. segregation) | *PPA2* | HGNC:28883 | NM_176869.3:c.833T>C p.(Leu278Ser) het 4<br>NM_176869.3:c.514G>A p.(Glu172Lys) het 5 | AR | Yes | HP:0000508 Ptosis; HP:0000602 Ophthalmoplegia; HP:0001337 Tremor; HP:0009830 Peripheral neuropathy; HP:0001638 Cardiomyopathy; | No | | PMID: 34400813 |
| 59 | P59 | Female | ES | adult | 5 | No | undiagnosed | unsolved | | | | | | HP:0003326 Myalgia; HP:0003236 Elevated creatine kinase ; HP:0002401 Stroke-like episode ; HP:0002076 Migraine ; HP:0000821 Hypothyroidism; | | | |
| 60 | P60 | Male | GS | adult | 8 | No | undiagnosed | unsolved | | | | | | HP:0000508 Ptosis; HP:0000602 Ophthalmoplegia; HP:0003324 Generalized muscle weakness; HP:0003236 Elevated creatine kinase ; HP:0001250 Seizure; | | | |
| 61 | P61 | Male | ES | adult | 10 | No | undiagnosed | unsolved | | | | | | HP:0000508 Ptosis; HP:0000602 Ophthalmoplegia; HP:0000407 Sensorineural hearing impairment ; HP:0000510 Retinitis pigmentosa; HP:0000819 Diabetes mellitus; | | | |
| 62 | P62 | Female | ES | pediatric | 9 | Yes | positive | solved by functional testing | *PDHA1* | HGNC:8806 | NM_000284.4:c.759+44G>A het 4 | XL | Yes | HP:0001252 Hypotonia; HP:0001257 Spasticity; HP:0001250 Seizure; HP:0002500 Abnormal cerebral white matter morphology ; HP:0000252 Microcephaly; | Yes | cDNA studies ( muscle) | |
| 63 | P63 | Male | ES | adult | 5 | No | undiagnosed | unsolved | | | | | | HP:0001324 Muscle weakness; HP:0003690 Limb muscle weakness; HP:0003200Ragged-red muscle fibers; HP:0003688 Cytochrome C oxidase-negative muscle fibers | | | |
| 64 | P64 | Female | ES | adult | 7 | No | undiagnosed | unsolved | | | | | | HP:0003324 Generalized muscle weakness; HP:0001252 Hypotonia ; HP:0009830 Peripheral neuropathy; HP:0000819 Diabetes mellitus; HP:0002666 Pheochromocytoma; | | | |
| 65 | P65 | Female | ES | adult | 5 | No | undiagnosed | unsolved | | | | | | HP:0001324 Muscle weakness; HP:0002076 Migraine ; HP:0001250 Seizure; HP:0009830 Peripheral neuropathy; HP:0000819 Diabetes mellitus; | | | |
| 66 | P66 | Female | GS | adult | 5 | No | undiagnosed | unsolved | | | | | | HP:0001324 Muscle weakness; HP:0001250 Seizure; HP:0000819 Diabetes mellitus; HP:0001410 Hepatopathy; HP:0002151 Increased serum lactate; | | | |
| 67 | P67 | Male | ES | pediatric | 8 | Yes | strong candidate | strong candidate - work in progress | *UQCRC1* | HGNC:12585 | NM_003365.3:c.1214-1G>T het<br>NM_003365.3:c.707-186G>A het | AR | Yes | HP:0000508 Ptosis; HP:0001252 Hypotonia ; HP:0001263 Global developmental delay; HP:0001250 Seizure; HP:0100704 Cerebral visual impairment; | Pending | cDNA studies | |
| 68 | P68 | Female | ES | pediatric | 5 | No | undiagnosed | unsolved | | | | | | HP:0001508 Failure to thrive ; HP:0002020 Gastro-oesophageal reflux; HP:0012211 Renal impairment; HP:0002151 Increased serum lactate | | | |
| 69 | P69 | Male | GS | pediatric | 7 | Yes | positive | solved by functional testing | *ECHS1* | HGNC:3151 | NM_004092.4:c.744T>G p.(Phe248Leu) het 4<br>NM_004092.4:c.489G>A p.(Pro163=) het 4 | AR | Yes | HP:0000508 Ptosis; HP:0001332 Dystonia; HP:0002376 Developmental regression; HP:0001250 Seizure; HP:0012751 Abnormal basal ganglia MRI signal intensity ; | Yes | Western blot and cDNA studies | |
| 70 | P70 | Male | ES | pediatric | 7 | No | undiagnosed | unsolved | | | | | | HP:0003324 Generalized muscle weakness; HP:0003546 Exercise intolerance; HP:0002076 Migraine ; HP:0002572 Episodic vomiting ; HP:0002151 Increased serum lactate; | | | |
| 71 | P71 | Female | GS | adult | 4 | No | undiagnosed | unsolved | | | | | | HP:0000508 Ptosis; HP:0000602 Ophthalmoplegia; HP:0001324 Muscle weakness; HP:0000407 Sensorineural hearing impairment ; HP:0004322 Short stature ; | | | |
| 72 | P72 | Male | ES | pediatric | 10 | Yes | positive | solved by functional testing | *ATAD3A*<br>*ATAD3B*<br>*ATAD3C* | HGNC:25567<br>HGNC:24007<br>HGNC:32151 | NC_000001.11:g.1456616-1524663dup het 5 | AD | Yes | HP:0007957 Corneal opacity; HP:0000518 Cataracts; HP:0001639 Hypertrophic cardiomyopathy ; HP:0001789 Hydrops fetalis ; HP:0002151 Increased serum lactate; | Yes | Proteomics, cDNA studies, qRT-PCR | PMID: 33575671 |
| 73 | P73 | Female | GS | adult | 5 | No | undiagnosed | unsolved | | | | | | HP:0000508 Ptosis; HP:0000602 Ophthalmoplegia; HP:0001324 Muscle weakness; HP:0000819 Diabetes mellitus; HP:0002151 Increased serum lactate; | | | |
| 74 | P74 | Male | GS | pediatric | 7 | No | undiagnosed | unsolved | | | | | | HP:0001252 Hypotonia ; HP:0001263 Global developmental delay; HP:0001508 Failure to thrive ;HP:0000122 Unilateral renal agenesis; HP:0002151 Increased serum lactate; | | | |
| 75 | P75 | Female | GS | adult | 7 | Yes | positive | solved by initial analysis (GS/ESmtDNAseq incl. segregation) | *POLG* | HGNC:9179 | NM_002693.3:c.2209G>C p.(Gly737Arg) het 5<br>NM_002693.3:c.695G>A p.(Arg232His) het 5 | AR | Yes | HP:0001324 Muscle weakness; HP:0002076 Migraine ; HP:0001250 Seizure; HP:0009830 Peripheral neuropathy; HP:0100704 Cerebral visual impairment | No | | |
| 76 | P76 | Female | GS | pediatric | 8 | Yes | positive | solved by initial analysis (GS/ESmtDNAseq incl. segregation) | *MT-ATP6* | HGNC:7414 | NC_012920.1:m.8611dup p.(Leu29Profs) hetp (62%) 5 | Mitochondrial | Yes | HP:0001252 Hypotonia ; HP:0001251 Ataxia; HP:0001249 Intellectual disability ; HP:0002151 Increased serum lactate; HP:0003535 3-methylglutaconic aciduria; | No | | |
| 77 | P77 | Female | GS | pediatric | 8 | Yes | positive | solved by initial analysis (GS/ESmtDNAseq incl. segregation) | *SLC19A3* | HGNC:16266 | NM_025243.4:c.337T>C p.(Tyr113His) het 5<br>NM_025243.4:c.223G>A p.(Asp75Asn) het 4 | AR | Yes | HP:0003324 Generalized muscle weakness; HP:0001332 Dystonia; HP:0001249 Intellectual disability ; HP:0001250 Seizure; HP:0012751 Abnormal basal ganglia MRI signal intensity; | No | | |
| 78 | P78 | Female | ES | pediatric | 7 | Yes | positive | solved by initial analysis (GS/ESmtDNAseq incl. segregation) | *PDHA1* | HGNC:8806 | NM_000284.4:c.905G>A p.(Arg302His) het 4 | XL | Yes | HP:0001257 Spasticity; HP:0001249 Intellectual disability ; HP:0002151 Increased serum lactate; HP:0003348 Hyperalaninemia; HP:0000252 Microcephaly | No | | |
| 79 | P79 | Male | GS | pediatric | 8 | Yes | positive | solved by functional testing | *ATAD3A*<br>*ATAD3B*<br>*ATAD3C* | HGNC:25567<br>HGNC:24007<br>HGNC:32151 | NC_000001.11:g.1456914_1524961dup het 5 | AD | Yes | HP:0001250 Seizure; HP:0007957 Corneal opacity; HP:0001639 Hypertrophic cardiomyopathy ; HP:0002151 Increased serum lactate; HP:0030890 White matter hyperintensities; | No | | PMID: 33575671 |
| 80 | P80 | Female | GS | adult | 5 | No | undiagnosed | unsolved | | | | | | HP:0000602 Ophthalmoplegia; HP:0001324 Muscle weakness; HP:0003326 Myalgia; HP:0003236 Elevated creatine kinase ; HP:0002076 Migraine ; | | | |
| 81 | P81 | Female | GS | adult | 5 | No | undiagnosed | unsolved | | | | | | HP:0000508 Ptosis; HP:0000602 Ophthalmoplegia; HP:0003324 Generalized muscle weakness; HP:0003326 Myalgia; HP:0003236 Elevated creatine kinase ; | | | |
| 82 | P82 | Female | GS | pediatric | 7 | Yes | positive | solved by initial analysis (GS/ESmtDNAseq incl. segregation) | *mtDNA deletion* | | NC_012920.1:m.5965_14996del hetp (50%) 5 | Mitochondrial | Yes | HP:0000508 Ptosis; HP:0000602 Ophthalmoplegia; HP:0000407 Sensorineural hearing impairment ;HP:0000819 Diabetes mellitus; HP:0012751 Abnormal basal ganglia MRI signal intensity | No | | |
| 83 | P83 | Female | GS | pediatric | 7 | Yes | positive | solved by reanalysis of GS or ES | *MORC2* | HGNC:23573 | NM_001303256.3:c.79G>A p.(Glu27Lys) het 5 | AD | No | HP:0001263 Global developmental delay; HP:0002151 Increased serum lactate; HP:0003348 Hyperalaninemia; HP:0012751 Abnormal basal ganglia MRI signal intensity ; HP:0033725 Thin corpus callosum | No | | |
| 84 | P84 | Male | GS | pediatric | 7 | Yes | strong candidate | strong candidate - work in progress | *EARS2* | HGNC:29419 | NM_001083614.2:c.845G>A p.(Gly282Asp) hom 3a | AR | Yes | HP:0001252 Hypotonia ; HP:0001332 Dystonia; HP:0001263 Global developmental delay; HP:0002151 Increased serum lactate; HP:0012747 Abnormal brainstem MRI signal intensity; | Pending | Western Blot and dipsticks (proband), affected sibiling 5-19% cox negative fibres proteomics and/or methylation testing pending | |
| 85 | P85 | Female | ES | pediatric | 6 | Yes | positive | solved by initial analysis (GS/ESmtDNAseq incl. segregation) | *ACAD9* | HGNC:21497 | NM_014049.5:c.796C>T p.(Arg266Trp) het 4<br>NM_014049.5:c.1429C>T p.(Arg477Ter) het 4 | AR | Yes | HP:0001263 Global developmental delay; HP:0001250 Seizure; HP:0002151 Increased serum lactate; HP:0003348 Hyperalaninemia; HP:0003535 3-methylglutaconic aciduria; | No | | |
| 86 | P86 | Male | GS | pediatric | 7 | Yes | positive | solved by initial analysis (GS/ESmtDNAseq incl. segregation) | *FBXL4* | HGNC:13601 | NM_001278716.2:c.486T>A p.(Tyr162Ter) het 5<br>NM_001278716.2:c.1444C>T p.(Arg482Trp) het 5 | AR | Yes | HP:0001251 Ataxia; HP:0001263 Global developmental delay; HP:0040289 Cyclic neutropenia ; HP:0002151 Increased serum lactate; HP:0012751 Abnormal basal ganglia MRI signal intensity | No | | |
| 87 | P87 | Female | ES | adult | 7 | No | undiagnosed | unsolved | | | | | | HP:0001324 Muscle weakness; HP:0003546 Exercise intolerance; HP:0003236 Elevated creatine kinase ; HP:0009830 Peripheral neuropathy; HP:0002151 Increased serum lactate | | | |
| 88 | P88 | Male | ES | pediatric | 6 | No | undiagnosed | unsolved | | | | | | HP:0001266 Choreoathetosis; HP:0001263 Global developmental delay; HP:0000407 Sensorineural hearing impairment ; HP:0000821 Hypothyroidism; HP:0012751 Abnormal basal ganglia MRI signal intensity ; | | | |
| 89 | P89 | Female | ES | pediatric | 5 | Yes | strong candidate | strong candidate - work in progress | *MORC2* | HGNC:23573 | NM_001303256.3:c.674T>A p.(Met225Lys) het 3a | AD | No | HP:0001251 Ataxia; HP:0001263 Global developmental delay; HP:0000407 Sensorineural hearing impairment ; HP:0000510 Retinitis pigmentosa; HP:0002500 Abnormal cerebral white matter morphology; | No | | |
| 90 | P90 | Female | ES | adult | 5 | No | undiagnosed | unsolved | | | | | | HP:0000508 Ptosis; HP:0000602 Ophthalmoplegia; HP:0001324 Muscle weakness; HP:0003546 Exercise intolerance; HP:0000407 Sensorineural hearing impairment ; | | | |
| 91 | P91 | Female | GS | pediatric | 7 | Yes | positive | solved by functional testing | *ECHS1* | HGNC:3151 | NM_004092.4:c.463G>A p.(Gly155Ser) het 5<br>NM_004092.4:c.796A>G p.(Thr266Ala) het 4 | AR | Yes | HP:0001249 Intellectual disability ; HP:0002376 Developmental regression; HP:0002151 Increased serum lactate; HP:0003348 Hyperalaninemia; HP:0012751 Abnormal basal ganglia MRI signal intensity | Yes | Reduced activity of short-chain enoyl-CoA hydratase consistent with ECHS1 deficiency (Fibroblasts) | |

| | | | | | | | | | | | | | | | | | |
|---|---|---|---|---|---|---|---|---|---|---|---|---|---|---|---|---|---|
| 92 | P92 | Female | ES | pediatric | 7 | Yes | positive | solved by reanalysis of GS or ES | *NDUFAF6* | HGNC:28625 | NM_152416.4:c.420+784C>T hom 5 | AR | Yes | HP:0001332 Dystonia; HP:0001337 Tremor; HP:0001250 Seizure; HP:0002151 Increased serum lactate; HP:0012751 Abnormal basal ganglia MRI signal intensity ; | No | | |
| 93 | P93 | Female | ES | adult | 4 | No | undiagnosed | unsolved | | | | | | HP:0003324 Generalized muscle weakness; HP:0001251 Ataxia; HP:0001249 Intellectual disability; HP:0012747Abnormal brainstem MRI signal intensity; HP:0002500 Abnormal cerebral white matter morphology ; | | | |
| 94 | P94 | Male | GS | adult | 4 | No | undiagnosed | unsolved | | | | | | HP:0003324 Generalized muscle weakness; HP:0003546 Exercise intolerance; HP:0004389 Intestinal pseudo-obstruction ; HP:0001733 Pancreatitis; HP:0002907 Microscopic hematuria | | | |
| 95 | P95 | Male | ES | adult | 9 | Yes | positive | solved by initial analysis (GS/ESmtDNAseq incl. segregation) | *POLG* | HGNC:9179 | NM_002693.3:c.2794C>T p.(His932Tyr) het 5<br>NM_002693.3:c.[752C>T;1760C>T] p.[(Thr251Ile)Pro587Leu)] het 5 | AR | Yes | HP:0000508 Ptosis; HP:0000602 Ophthalmoplegia; HP:0001324 Muscle weakness; HP:0009830 Peripheral neuropathy; HP:0007957 Corneal opacity; | No | | |
| 96 | P96 | Male | ES | adult | 6 | No | undiagnosed | unsolved | | | | | | HP:0002401 Stroke-like episode ; HP:0002076 Migraine ; HP:0001250 Seizure; HP:0000407 Sensorineural hearing impairment | | | |
| 97 | P97 | Female | ES | pediatric | 5 | Yes | strong candidate | strong candidate - work in progress | *NUS1* | HGNC:21042 | NM_138459.5:c.865C>T p.(Gln289Ter) het 3a | AD | No | HP:0003324 Generalized muscle weakness; HP:0001337 Tremor; HP:0100660 Dyskinesia ; HP:0001249 Intellectual disability ; HP:0002151 Increased serum lactate; | No | | |
| 98 | P98 | Female | GS | pediatric | 5 | Yes | positive | solved by initial analysis (GS/ESmtDNAseq incl. segregation) | *TRNT1* | HGNC:17341 | NM_182916.3:c.218_219insGAGGGATTTATTAAATTATAAATTTATTAAATGGATTTATTAAA p.(Lys74ArgfsTer14) het 4<br>NM_182916.3:c.1246A>G p.(Lys416Glu) het 4 | AR | Yes | HP:0009830 Peripheral neuropathy; HP:0000407 Sensorineural hearing impairment ; HP:0000648 Optic atrophy ; HP:0004322 Short stature ; HP:0001903 Anaemia; | No | | |
| 99 | P99 | Male | GS | pediatric | 10 | Yes | positive | solved by initial analysis (GS/ESmtDNAseq incl. segregation) | *AARS2* | HGNC:21022 | NM_020745.4:c.1774C>T p.(Arg592Trp) het 5<br>NM_020745.4:c.1528A>G p.(Thr510Ala) het 4 | AR | Yes | HP:0001639 Hypertrophic cardiomyopathy ; HP:0002151 Increased serum lactate; HP:0003688 Cytochrome C oxidase-negative muscle fibers; HP:0008322 Abnormal mitochondrial morphology ; HP:0008972 Decreased activity of mitochondrial respiratory chain | No | | |
| 100 | P100 | Female | ES | adult | 5 | No | undiagnosed | unsolved | | | | | | HP:0001324 Muscle weakness; HP:0003546 Exercise intolerance; HP:0003236 Elevated creatine kinase ; HP:0002015 Dysphagia; HP:0002076 Migraine ; | | | |
| 101 | P101 | Female | ES | pediatric | 6 | Yes | strong candidate | strong candidate - work in progress | *VARS1* | HGNC:12651 | NM_006295.3:c.1609C>T p.(Arg537Trp) het 4<br>NM_006295.3:c.1031C>A p.(Pro344His) het 3a | AR | No | HP:0003324 Generalized muscle weakness; HP:0001252 Hypotonia ; HP:0001263 Global developmental delay; HP:0002151 Increased serum lactate; HP:0000252 Microcephaly | Pending | VARS1 functional studies | |
| 102 | P102 | Female | ES | pediatric | 6 | Yes | positive | solved by initial analysis (GS/ESmtDNAseq incl. segregation) | *PPM1D* | HGNC:9277 | NM_003620.4:c.1451T>A p.(Leu484Ter) het 4 | AD | No | HP:0001252 Hypotonia ; HP:0001263 Global developmental delay; HP:0002076 Migraine ; HP:0001250 Seizure; HP:0002151 Increased serum lactate; | No | | |
| 103 | P103 | Female | ES | pediatric | 5 | No | undiagnosed | unsolved | | | | | | HP:0003324 Generalized muscle weakness; HP:0003326 Myalgia; HP:0009830 Peripheral neuropathy; HP:0001638 Cardiomyopathy; HP:0002151 Increased serum lactate; | | | |
| 104 | P104 | Female | GS | pediatric | 5 | Yes | positive | solved by reanalysis of GS or ES | *BPTF* | HGNC:3581 | NM_182641.4:c.8645dup p.(Ser2883LysfsTer2) het 4 | AD | No | HP:0000508 Ptosis; HP:0000602 Ophthalmoplegia; HP:0001324 Muscle weakness; HP:0000819 Diabetes mellitus; HP:0004322 Short stature ; | No | | |
| 105 | P105 | Male | GS | pediatric | 4 | Yes | positive | solved by reanalysis of GS or ES | *ZFYVE19* | HGNC:20758 | NM_001077268.2:c.314C>G p.(Ser105Ter) hom 5 | AR | No | HP:0001263 Global developmental delay; HP:0000407 Sensorineural hearing impairment; HP:0002240 Hepatomegaly; HP:0001396 Cholestasis ; HP:0001903 Anaemia; | No | | |
| 106 | P106 | Female | ES | pediatric | 6 | Yes | positive | solved by initial analysis (GS/ESmtDNAseq incl. segregation) | *KCNJ10* | HGNC:6256 | NM_002241.5:c.475del p.(Ile159SerfsTer39) het 4<br>NM_002241.5:c.68del p.(Pro23GlnfsTer9) het 4 | AR | No | HP:0001266 Choreoathetosis; HP:0001249 Intellectual disability ; HP:0001250 Seizure; HP:0000407 Sensorineural hearing impairment ; HP:0012751 Abnormal basal ganglia MRI signal intensity ; | No | | |
| 107 | P107 | Female | GS | pediatric | 6 | Yes | positive | solved by initial analysis (GS/ESmtDNAseq incl. segregation) | *JAM3* | HGNC:15532 | NM_032801.5:c.1A>G p.(Met1?) hom 4 | AR | No | HP:0001332 Dystonia; HP:0001263 Global developmental delay; HP:0001250 Seizure; HP:0000518 Cataracts; HP:0000238 Hydrocephalus | No | | |
| 108 | P108 | Female | GS | pediatric | 6 | Yes | positive | solved by initial analysis (GS/ESmtDNAseq incl. segregation) | *POLG* | HGNC:9179 | NM_002693.3:c.911T>G p.(Leu304Arg) hom 5 | AR | Yes | HP:0000508 Ptosis; HP:0000602 Ophthalmoplegia; HP:0003324 Generalized muscle weakness; HP:0003546 Exercise intolerance;HP:0009830 Peripheral neuropathy | No | | |
| 109 | P109 | Female | ES | adult | 4 | No | undiagnosed | unsolved | | | | | | HP:0000508 Ptosis; HP:0000602 Ophthalmoplegia; HP:0003324 Generalized muscle weakness; HP:0003326 Myalgia; HP:0000407 Sensorineural hearing impairment | | | |
| 110 | P110 | Male | GS | pediatric | 6 | Yes | positive | solved by initial analysis (GS/ESmtDNAseq incl. segregation) | *MT-ATP6* | HGNC:7414 | NC_012920.1:m.8969G>A p.(Ser148Asn) hetp (85%) 5 | Mitochondrial | Yes | HP:0003546 Exercise intolerance; HP:0001263 Global developmental delay; HP:0001639 Hypertrophic cardiomyopathy ; HP:0001903 Anaemia; HP:0002151 Increased serum lactate; | No | | |
| 111 | P111 | Male | GS | pediatric | 8 | Yes | strong candidate | strong candidate - work in progress | *MT-ND6* | HGNC:7462 | NC_012920.1:m.14594T>C p.(Tyr27Cys) hetp (49%) 3a | Mitochondrial | Yes | HP:0001324 Muscle weakness; HP:0001266 Choreoathetosis; HP:0001249 Intellectual disability ; HP:0002151 Increased serum lactate; HP:0012751 Abnormal basal ganglia MRI signal intensity ; | Pending | Proteomics BN-PAGE | |
| 112 | P112 | Female | GS | adult | 5 | No | undiagnosed | unsolved | | | | | | HP:0001324 Muscle weakness; HP:0001283 Bulbar palsy;HP:0002076 Migraine ; HP:0001250 Seizure; HP:0000407 Sensorineural hearing impairment ; | | | |
| 113 | P113 | Female | GS | adult | 6 | No | undiagnosed | unsolved | | | | | | HP:0000508 Ptosis; HP:0003326 Myalgia; HP:0003546 Exercise intolerance; HP:0000360 Tinnitus ; HP:0002019 Constipation; | | | |
| 114 | P114 | Female | ES | adult | 5 | No | undiagnosed | unsolved | | | | | | HP:0001324 Muscle weakness;HP:0003546 Exercise intolerance; HP:0002376 Developmental regression; HP:0002076 Migraine ; HP:0002151 Increased serum lactate; | | | |
| 115 | P115 | Male | ES | pediatric | 6 | Yes | positive | solved by reanalysis of GS or ES | *CTBP1* | HGNC:2494 | NM_001012614.2:c.991C>Tp.(Arg331Trp) het 5 | AD | Yes | HP:0003324 Generalized muscle weakness; HP:0002376 Developmental regression; HP:0009830 Peripheral neuropathy; HP:0000407 Sensorineural hearing impairment; HP:0001272 Cerebellar atrophy | No | | |
| 116 | P116 | Female | GS | adult | 7 | No | undiagnosed | unsolved | | | | | | HP:0000508 Ptosis; HP:0000602 Ophthalmoplegia; HP:0001324 Muscle weakness; HP:0000518 Cataracts; HP:0000819 Diabetes mellitus; | | | |
| 117 | P117 | Female | ES | pediatric | 6 | Yes | partial | solved by reanalysis of GS or ES | *MYH7* | HGNC:7577 | NM_000257.4:c.428G>A p.(Arg143Gln) het 4 | AD | No | HP:0001639 Hypertrophic cardiomyopathy ; HP:0001266 Choreoathetosis; HP:0001250 Seizure; HP:0004322 Short stature ; HP:0001733 Pancreatitis; | No | | |
| 118 | P118 | Female | ES | pediatric | 5 | No | undiagnosed | unsolved | | | | | | HP:0003326 Myalgia; HP:0003236 Elevated creatine kinase ; HP:0000822 Hypertension; HP:0012211 Renal impairment; HP:0002151 Increased serum lactate | | | |
| 119 | P119 | Male | GS | adult | 4 | Yes | positive | solved by initial analysis (GS/ESmtDNAseq incl. segregation) | *MYH9 , USH2A* | HGNC:7579, HGNC:12601 | NM_002473.6:c.2114G>A p.(Arg705His) het 5<br>NM_206933.4:c.14453C>T p.(Pro4818Leu) het 4 | AD | No | HP:0001252 Hypotonia ; HP:0000407 Sensorineural hearing impairment ; HP:0000510 Retinitis pigmentosa; HP:0000819 Diabetes mellitus; HP:0012211 Renal impairment; | No | | PMID: 35128751 |
| 120 | P120 | Male | GS | adult | 4 | No | undiagnosed | unsolved | | | | | | HP:0000508 Ptosis; HP:0000602 Ophthalmoplegia;HP:0000819 Diabetes mellitus; HP:0000135 Hypogonadism; HP:0002151 Increased serum lactate | | | |
| 121 | P121 | Female | GS | adult | 4 | No | undiagnosed | unsolved | | | | | | HP:0000508 Ptosis; HP:0000602 Ophthalmoplegia;HP:0001251 Ataxia; HP:0000407 Sensorineural hearing impairment ; HP:0002500 Abnormal cerebral white matter morphology | | | |
| 122 | P122 | Female | ES | adult | 6 | No | undiagnosed | unsolved | | | | | | HP:0000508 Ptosis; HP:0000602 Ophthalmoplegia; HP:0003546 Exercise intolerance; HP:0003200Ragged-red muscle fibers; HP:0003688 Cytochrome C oxidase-negative muscle fibers | | | |
| 123 | P123 | Female | GS | pediatric | 7 | No | undiagnosed | unsolved | | | | | | HP:0003324 Generalized muscle weakness; HP:0001332 Dystonia; HP:0001263 Global developmental delay; HP:0001250 Seizure; HP:0000407 Sensorineural hearing impairment ; | | | |
| 124 | P124 | Female | ES | pediatric | 8 | Yes | positive | solved by muscle mtDNA testing | *mtDNA deletion* | | NC_012920.1:m.7821_14798del hetp (% not quantified) 5 | Mitochondrial | Yes | HP:0000508 Ptosis; HP:0000602 Ophthalmoplegia; HP:0001324 Muscle weakness; HP:0000407 Sensorineural hearing impairment; HP:0012751 Abnormal basal ganglia MRI signal intensity ; | No | | |
| 125 | P125 | Male | GS | pediatric | 6 | Yes | strong candidate | strong candidate - work in progress | *MT-ATP6* | HGNC:7414 | NC_012920.1:m.8812A>T p.(Thr96Ser) homp 3a | Mitochondrial | Yes | HP:0003324 Generalized muscle weakness; HP:0001252 Hypotonia ; HP:0001257 Spasticity; HP:0001263 Global developmental delay; HP:0001249 Intellectual disability ; HP:0001250 Seizure; HP:0001508 Failure to thrive ; HP:0002019 Constipation; HP:0002151 Increased serum lactate; HP:0002352 Leukoencephalopathy ; HP:0030890 White matter hyperintensities | Pending | proteomics | |
| 126 | P126 | Male | ES | pediatric | 8 | No | undiagnosed | unsolved | | | | | | HP:0003236 Elevated creatine kinase ; HP:0001250 Seizure; HP:0000821 Hypothyroidism; HP:0000846 Adrenal insufficiency; HP:0012751 Abnormal basal ganglia MRI signal intensity ; | | | |
| 127 | P127 | Male | ES | pediatric | 7 | No | undiagnosed | unsolved | | | | | | HP:0000508 Ptosis; HP:0003324 Generalized muscle weakness; HP:0001250 Seizure; HP:0009830 Peripheral neuropathy; HP:0002500 Abnormal cerebral white matter morphology ; | | | |
| 128 | P128 | Male | ES | pediatric | 5 | Yes | positive | solved by functional testing | *PNPT1* | HGNC:23166 | NM_033109.5:c.2212C>T p.(Arg738Cys) het 5<br>NM_033109.5:c.2050G>A p.(Ala684Thr) het 5 | AR | Yes | HP:0000508 Ptosis; HP:0000602 Ophthalmoplegia; HP:0001324 Muscle weakness; HP:0001251 Ataxia; HP:0000407 Sensorineural hearing impairment ; | Yes | Western blot and qRT-PCR | PMID: 31752325 |
| 129 | P129 | Female | GS | adult | 7 | No | undiagnosed | unsolved | | | | | | HP:0000508 Ptosis; HP:0003324 Generalized muscle weakness; HP:0003236 Elevated creatine kinase ; HP:0002076 Migraine ; HP:0002151 Increased serum lactate; | | | |
| 130 | P130 | Female | ES | pediatric | 6 | Yes | positive | solved by functional testing | *MT-ATP6* | HGNC:7414 | NC_012920.1:m.8672T>C hetp (81%) 4 | Mitochondrial | Yes | HP:0001257 Spasticity; HP:0001249 Intellectual disability ; HP:0001638 Cardiomyopathy; HP:0002151 Increased serum lactate; HP:0003535 3-methylglutaconic aciduria; | Yes | Proteomics | |
| 131 | P131 | Female | GS | pediatric | 6 | No | undiagnosed | unsolved | | | | | | HP:0001638 Cardiomyopathy; HP:0001639 Hypertrophic cardiomyopathy ; HP:0002151 Increased serum lactate; HP:0003348 Hyperalaninemia | | | |
| 132 | P132 | Male | ES | pediatric | 5 | No | undiagnosed | unsolved | | | | | | HP:0001252 Hypotonia ; HP:0001332 Dystonia; HP:0000407 Sensorineural hearing impairment ; HP:0012751 Abnormal basal ganglia MRI signal intensity ; HP:0002453Abnormal globus pallidus morphology ; | | | |
| 133 | P133 | Female | ES | pediatric | 5 | Yes | strong candidate | strong candidate - work in progress | *UNC13A* | HGNC:23150 | NM_001080421.3:c.2423G>A p.(Gly808Asp) het 3a | AD | No | HP:0001252 Hypotonia ; HP:0001266 Choreoathetosis; HP:0001249 Intellectual disability ; HP:0001250 Seizure; HP:0002151 Increased serum lactate; | Pending | | |
| 134 | P134 | Male | GS | pediatric | 5 | Yes | positive | solved by initial analysis (GS/ESmtDNAseq incl. segregation) | *GFAP* | HGNC:4235 | NM_002055.5:c.236G>A p.(Arg79His) het 5 | AD | No | HP:0003324 Generalized muscle weakness; HP:0001252 Hypotonia ; HP:0002019 Constipation; HP:0002500 Abnormal cerebral white matter morphology | No | | |
| 135 | P135 | Male | GS | pediatric | 8 | Yes | positive | solved by functional testing | *AARS2* | HGNC:21022 | NM_020745.4:c.2870C>T p.(Ser957Leu) het 4<br>NC_000006.11:g.44274355_44278496del het 5 | AR | Yes | HP:0003236 Elevated creatine kinase ; HP:0001263 Global developmental delay; HP:0002151 Increased serum lactate; HP:0500233 Increased CSF alanine concentration ; HP:0025045 Abnormal brain lactate level by MRS; | Yes | Western blot and cDNA studies | PMID: 33924034 |
| 136 | P136 | Female | GS | pediatric | 5 | Yes | positive | solved by functional testing | *MRPS34* | HGNC:16618 | NM_023936.2:c.94C>T p.(Gln32Ter) het 5<br>NM_023936.2:c.598_605del p.(Pro200LeufsTer36) het 4 | AR | Yes | HP:0000602 Ophthalmoplegia; HP:0003324 Generalized muscle weakness; HP:0001249 Intellectual disability ; HP:0002151 Increased serum lactate; HP:0012751 Abnormal basal ganglia MRI signal intensity ; | Yes | Western blot and cDNA studies | |
| 137 | P137 | Male | GS | pediatric | 6 | No | undiagnosed | unsolved | | | | | | HP:0003324 Generalized muscle weakness; HP:0001252 Hypotonia ; HP:0001263 Global developmental delay; HP:0001396 Cholestasis ; HP:0002151 Increased serum lactate; | | | |
| 138 | P138 | Male | GS | adult | 6 | Yes | positive | solved by initial analysis (GS/ESmtDNAseq incl. segregation) | *MT-ND3* | HGNC:7458 | NC_012920.1:m.10191T>C p.(Ser45Pro) hetp (14%) 5 | Mitochondrial | Yes | HP:0000508 Ptosis; HP:0003324 Generalized muscle weakness; HP:0002401 Stroke-like episode; HP:0001250 Seizure; HP:0002151 Increased serum lactate; | No | | |
| 139 | P139 | Male | GS | pediatric | 8 | Yes | positive | solved by initial analysis (GS/ESmtDNAseq incl. segregation) | *SURF1* | HGNC:11474 | NM_003172.4:c.587A>G p.(Gln196Arg) het 4<br>NM_003172.4:c.312_321delinsAT p.(Leu105Ter) het 5 | AR | Yes | HP:0000602 Ophthalmoplegia; HP:0001332 Dystonia; HP:0001263 Global developmental delay; HP:0000510 Retinitis pigmentosa; HP:0012751 Abnormal basal ganglia MRI signal intensity; | No | | |
| 140 | P140 | Male | GS | pediatric | 8 | Yes | positive | solved by initial analysis (GS/ESmtDNAseq incl. segregation) | *MT-TS1* | HGNC:7497 | NC_012920.1:m.7512T>C homp 4 | Mitochondrial | Yes | HP:0001324 Muscle weakness; HP:0003236 Elevated creatine kinase ; HP:0001249 Intellectual disability ; HP:0000407 Sensorineural hearing impairment ; HP:0031546 Cardiac conduction abnormality ; | No | | |

ES Exome Sequencing; GS Genome Sequencing; MNC modified Nijmegen criteria; homp homoplasmic; hetp heteroplasmic; hom homozygous; het heterozygous; HGNC ID HUGO Gene Nomenclature Committee identifier ; AD Autosomal Dominant, AR Autosomal Recessive, XL X linked

* The table displays the 5 most relevant HPO terms per individual, the total number of HPO terms per individual ranged from 4 to 23.

Biochemical analyses prompted by our genomic findings further supported the pathogenicity of variants in non-mitochondrial disease genes already classified as Class 5:

+ DHCR7 (P31): Elevated plasma level of 7-dehydrocholesterol (126 µmol/L; normal range < 5 µmol/L) consistent with a deficiency of 7-dehydrocholesterol reductase.

++ AMACR (P23): Elevated plasma levels of pristanate 132 µmol/L (normal range 0-2.5) and phytanate 26 µmol/L (normal range 0-20 µmol/L) with a normal VLCFA profile (C24:C22 ratio of 0.771; normal range 0.55-1.05, and C26/C22 ratio of 0.013; normal range 0-0.03) consistent with a deficiency of alpha-methylacyl-CoA racemase.
