## Supplementary material for "The Australian Genomics Mitochondrial Flagship: A National Program Delivering Mitochondrial Diagnoses": Table S4

**Table S4. mtDNA variants identified from off-target WES**

| | | mtDNAseq blood result | Seen in WES in blood reanalysis |
|---|---|---|---|
| P11 | *MT-ND5* | NC_012920.1:m.13513G>A p.(Asp393Asn) hetp (64%) 5 | Yes, hetp 51% (read depth 131X) |
| P18 | *MT-TV* | NC_012920.1:m.1638T>C homp 4 | no |
| P25 | *MT-TW* | NC_012920.1:m.5559A>G homp 5 | no |
| P33 | *MT-ND3* | NC_012920.1:m.10191T>C p.(Ser45Pro) hetp (22%) 5 | no |
| P43 | *MT-ND1* | NC_012920.1:m.3697G>A p.(Gly131Ser) hetp (53%) 5 | Yes, hetp 52% (read depth 211X) |
| P130 | *MT-ATP6* | NC_012920.1:m.8672T>C hetp (81%) 4 | Yes, hetp (44%) (read depth 86X) |
