## Supplementary material for "The Australian Genomics Mitochondrial Flagship: A National Program Delivering Mitochondrial Diagnoses": Table S5

### Table S5. Likely Diagnosis between European, Non-European, and Unknown reported ancestries

#### Pediatric-onset

| | | European | (n39) | Non-European | (n41) | Unknown | (n5) |
|---|---|---|---|---|---|---|---|
| | | **n** | **%** | **n** | **%** | **n** | **%** |
| | **likely diagnosis** | 26 | 67% | 32 | 78% | 3 | 60% |
| mitochondrial disease gene | AD | 2 | 5% | 2 | 5% | | |
| | AR | 10 | 26% | 11 | 27% | 1 | 20% |
| | XL | 2 | 5% | 1 | 2% | | |
| | mtDNA | 4 | 10% | 8 | 20% | 1 | 20% |
| non-mitochondrial | AD | 7 | 18% | 4 | 5% | | |
| | AR | 1 | 3% | 6 | 5% | | |

#### Adult-onset

| | | European | (n43) | Non-European | (n4) | Unknown | (n8) |
|---|---|---|---|---|---|---|---|
| | | **n** | **%** | **n** | **%** | **n** | **%** |
| | **likely diagnosis** | 12 | 28% | 1 | 25% | 4 | 50% |
| mitochondrial disease gene | AD | | | | | 1 | 13% |
| | AR | 2 | 5% | | | | |
| | mtDNA | 5 | 12% | | | 2 | 25% |
| | mtDNA + AR | 2 | 5% | | | 1 | 13% |
| non-mitochondrial | AD | 2 | 5% | | | | |
| | AR | 1 | 2% | 1 | 25% | | |
